# Integration of genetic colocalizations with physiological and pharmacological perturbations identifies cardiometabolic disease genes

**DOI:** 10.1101/2021.09.28.21264208

**Authors:** Michael J. Gloudemans, Brunilda Balliu, Daniel Nachun, Matthew G. Durrant, Erik Ingelsson, Martin Wabitsch, Thomas Quertermous, Stephen B. Montgomery, Joshua W. Knowles, Ivan Carcamo-Orive

**Affiliations:** Biomedical Informatics Training Program, Stanford, CA, USA; Department of Pathology, Stanford, CA, USA; Department of Computational Medicine, UCLA, CA, USA; Department of Genetics, Stanford, CA, USA; Department of Immunology, Stanford, CA, USA; Department of Medicine, Division of Cardiovascular Medicine and Cardiovascular Institute, Stanford, CA, USA; Department of Pediatrics and Adolescent Medicine, Division of Pediatric Endocrinology, Ulm University, Ulm, Germany; Diabetes Research Center, Stanford, CA, USA; Prevention Research Center, Stanford, CA, USA

**Keywords:** genome-wide association studies, integrative gene prioritization, colocalization, differential expression, perturbation experiments, insulin resistance, type 2 diabetes

## Abstract

**Background:** Identification of causal genes for polygenic human diseases has been extremely challenging, and our understanding of how physiological and pharmacological stimuli modulate genetic risk at disease-associated loci is limited. Specifically, insulin resistance (IR), a common feature of cardiometabolic disease, including type 2 diabetes, obesity, and dyslipidemia, lacks well-powered GWAS, and therefore few associated loci and causal genes have been identified.

**Results:** Here, we perform and integrate LD-adjusted colocalization analyses across nine cardiometabolic traits combined with eQTLs and sQTLs from five metabolically relevant human tissues (subcutaneous and visceral adipose, skeletal muscle, liver, and pancreas). We identify 470 colocalized loci and prioritize 207 loci with a single colocalized gene. To elucidate upstream regulators and functional mechanisms for these genes, we integrate their transcriptional responses to 21 physiological and pharmacological cardiometabolic regulators in human adipocytes, hepatocytes, and skeletal muscle cells, and map their protein-protein interactions.

**Conclusions:** Our use of transcriptional responses under metabolic perturbations to contextualize genetic associations from our state-of-the-art colocalization approach provides a list of likely causal genes and their upstream regulators in the context of IR-associated cardiometabolic risk.

## Background

Cardiometabolic diseases linked with insulin resistance (IR), including type 2 diabetes (T2D), obesity, and dyslipidemia, have reached staggering prevalence and are major causes of morbidity and mortality. Genome-wide association studies (GWAS) have identified hundreds of loci containing thousands of candidate genes associated with these cardiometabolic diseases and have shown that they have partially overlapping genetic architectures. For instance, in the case of T2D, GWAS have identified over 300 distinct susceptibility loci [1, 2, 3, 4] that harbor thousands of genes. Recent work [5] has identified subgroups of individuals with differential risk for other cardiometabolic traits, e.g., fasting insulin, fasting glucose, waist-hip ratio (WHR), body mass index (BMI), triglycerides (TG), and high-density lipoprotein cholesterol (HDL), helping to account for the observed clinical heterogeneity in T2D. Thus, the combination of different polygenic risk pathways, including insulin action, insulin secretion, obesity, fat distribution, and lipids/liver function, forms an overall palette of risk [5, 6, 7, 8, 9]. These polygenic clusters highlight the close relationship between IR, T2D, and cardiometabolic traits and confirm the central role of peripheral tissues (adipose tissue, skeletal muscle, and liver) in IR [10].

While most T2D causal genes discovered so far are related to insulin production or secretion [11, 12, 13, 14, 15, 16], partly because GWAS for direct measures of insulin sensitivity have been small [17, 18], mounting evidence suggests that some T2D loci increase risk directly through IR [17, 19, 20, 21, 22], and many other loci have not yet been categorized. This knowledge gap has hampered therapeutic advances. However, large-scale GWAS of various cardiometabolic traits now provide a new opportunity for identifying cardiometabolic risk genes and partitioning them into IR and non-IR-related sets by jointly analyzing multiple intermediate traits for cardiometabolic disease.

Beyond genetic variation alone, risk genes for human disease are also modulated by various physiological and pharmacological stimuli whose biological effects are not yet fully characterized. In addition to fundamental stimuli like glucose and insulin, inflammatory cytokines such as TNF-*α* and IL-6 can also impair glycemic control and affect cardiometabolic disease-related transcriptional regulation [23]. In conjunction with these physiological factors, drugs used for the treatment of cardiometabolic diseases, such as atorvastatin, metformin, and rosiglitazone, modulate the activity of disease-relevant genetic pathways. A precise model of how these and other extrinsic factors affect cardiometabolic disease via intermediary genes is complex and still growing [23]. Thus, determining how these known upstream regulators modify transcription of risk genes will enhance our mechanistic understanding of cardiometabolic disease and IR biology.

To advance the identification and prioritization of causal genes for cardiometabolic traits and IR, and to shed light upon their functional contexts, we systematically integrate GWAS- and QTL-derived genetic signals with metabolic regulators. Specifically, we perform a state-of-the-art colocalization analysis on twelve publicly available GWAS comprising nine different IR and cardiometabolic traits, using eQTLs and sQTLs detected in five metabolically relevant tissues that directly impact glucose homeostasis: subcutaneous adipose and visceral adipose tissue, liver, skeletal muscle, and pancreas. We identify patterns of pancreatic and non-pancreatic tissue specificity and trait sharing at colocalized loci, establishing a knowledge-based priority list of uniquely colocalized candidate causal genes. To elucidate the functional mechanisms of these candidate causal genes, we integrate data on transcriptional responses to 21 cardiometabolically relevant perturbations in human adipocytes, hepatocytes, and skeletal muscle cells. Integrating these results, we annotate and prioritize 48 candidate cardiometabolic causal genes with association to IR or T2D, 64 with association to WHR, and 57 with association to TG or HDL. Our results enable fine-scale dissection of these candidates and prioritization of high-confidence cardiometabolic risk genes as potential therapeutic targets.

## Results

### Colocalization analysis associates GWAS traits with QTLs in disease-relevant tissues

To identify candidate causal genes for cardiometabolic disease, we first performed colocalization analysis of eQTLs and sQTLs in five human tissues across 9 cardiometabolic traits from 12 separate GWAS (Tables S1 & S2). Fig. S1 illustrates the process we used to select genome-wide significant loci and overlapping eQTL/sQTL features for colocalization testing. We first identified 2,859 independent SNP-trait associations (Additional Data S1). Since these traits can share causal variants, we binned each locus into one of 1,724 independent and previously defined partitions of the genome [24] (Additional Data S2). This assured that the mapping of associations to loci was invariant to the total number of GWAS traits. Of these 1,724 loci, 1,071 contained at least one GWAS association (Fig. S2) and were considered in subsequent analyses.

We identified all genes expressed in at least one of five relevant GTEx tissues (subcutaneous and visceral adipose, skeletal muscle, liver, and pancreas) with a transcriptional start site (TSS) less than 1 Mb from one or more GWAS lead variants, rendering a total of 22,105 candidate genes, including protein-coding genes, long non-coding RNAs, and other non-coding transcripts (Fig. 1A). We then filtered to the genes with at least one eQTL or sQTL (p *<* 1e-5) overlapping the lead GWAS SNP (Fig. S1), leaving 817 loci containing 4,704 candidate causal genes. Accordingly, 254 GWAS loci (24%) had no traceable eQTL or sQTL association and were excluded from subsequent analyses (Fig. S2).

**Figure 1:**
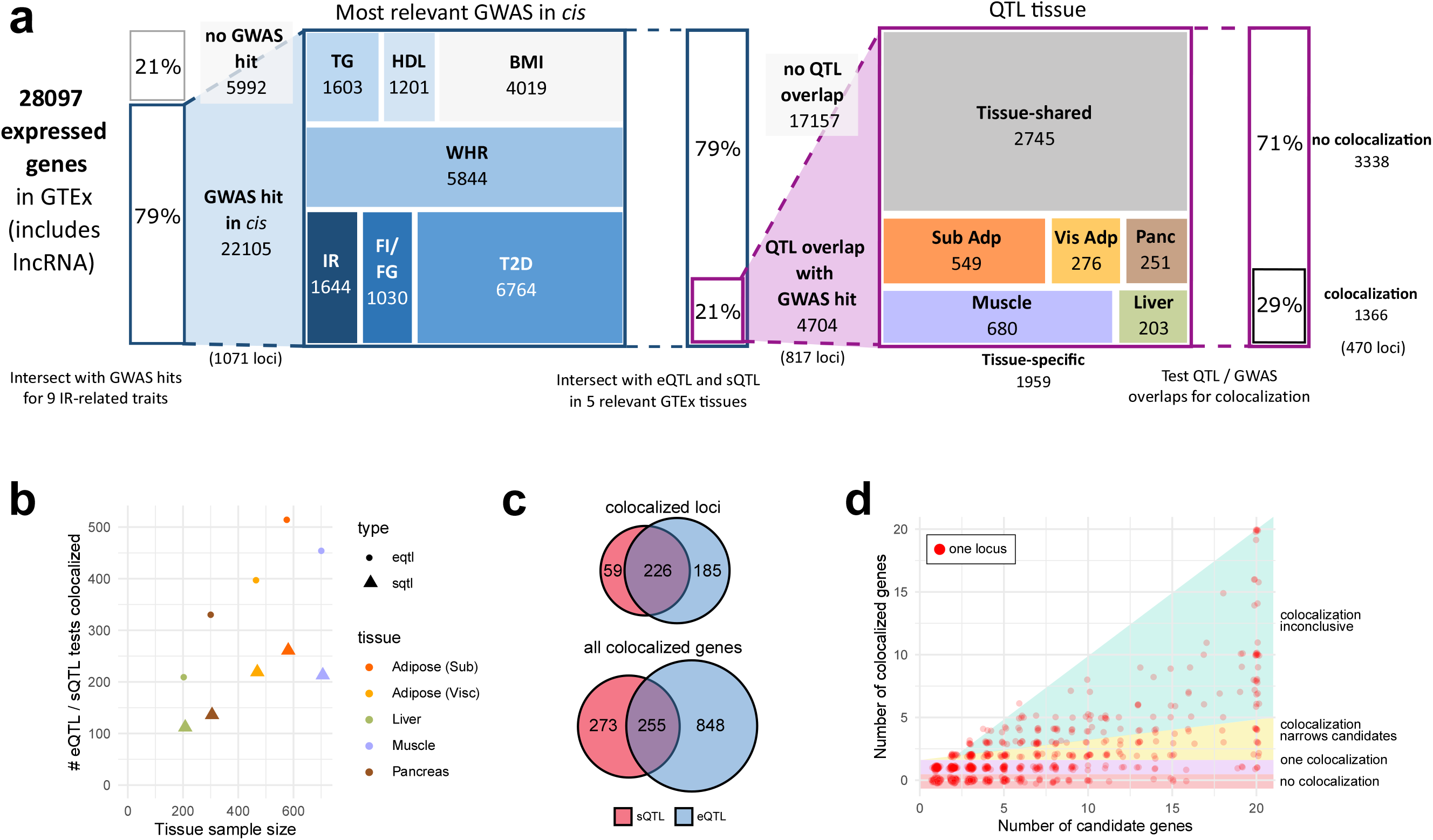
Colocalization testing narrows candidate genes across cardiometabolic traits, tissues, and QTL types. (A) Candidate genes are filtered based on GWAS proximity, GWAS / QTL overlap, and colocalization testing. Box sizes at each step are proportional to the relative abundance within each category. (B) Scatterplot of the relationship between sample size and number of eQTL / sQTL colocalizations in a given tissue. (C) Number of loci and number of individual genes with colocalizations for sQTLs, eQTLs, or both. (D) Scatterplot of the relationship between the number of candidates (number genes with QTL overlapping a lead GWAS variant) and number of colocalizations at each locus.

We performed colocalization analysis to identify loci with a common causal variant affecting both a cardiometabolic GWAS trait and a transcriptional QTL phenotype. To avoid sensitivity to local variation in LD structures, we implemented our own LD-modified adaptation of the canonical eCAVIAR colocalization analysis (Methods & Fig. S3). We observed colocalization for 470 (44%) of the 1,071 GWAS loci, across all QTL tissues and GWAS traits (Tables S3, S4, & Additional Data S3). The number of colocalized genes per tissue was correlated with tissue sample sizes in GTEx (*ρ* = 0.90, Fig. 1B). While in some instances both eQTL and sQTL colocalizations point to the same gene (Fig. 1C), 20% of colocalized genes would have not been detected without sQTLs. For example, the adipose-specific colocalization *BDNF-AS* showed sQTL but not eQTL colocalization. The lead cluster of candidate causal variants at this locus is located within the body of the antisense *BDNF-AS* gene, farther away from the *BDNF* gene and the *BDNF-AS* promoter. Our results underscore the advantage of colocalization analyses with both eQTL and sQTL variants.

### Disease loci harbor different causal architectures

The number of candidate genes within each locus (i.e., genes with a QTL overlapping a lead GWAS variant) and the number of colocalized genes varied extensively (Figs. 1D & 2A), as did the colocalized tissues and traits at these loci (Fig. 2B-C). To quantify the ability of colocalization analysis to narrow down the number of candidate causal genes within a locus, we classified loci according to the number of initial candidate genes and the number of colocalized genes (Fig. 2A). Of the 197 loci with a single candidate gene, just under a quarter (46 loci) colocalized, highlighting the utility of colocalization testing to inform functional follow-up, even at loci for which there is only one candidate gene with an overlapping QTL. In total, we identified 207 loci with a single colocalized gene (25% of all tested loci). In line with previous estimates [25], 41% of uniquely colocalized genes were the nearest gene to the lead GWAS SNP (Fig. S4). This percentage was even lower (17%) when looking at all colocalized genes. These results emphasize the value of colocalization analyses over approaches that assume the nearest gene to be causal.

**Figure 2:**
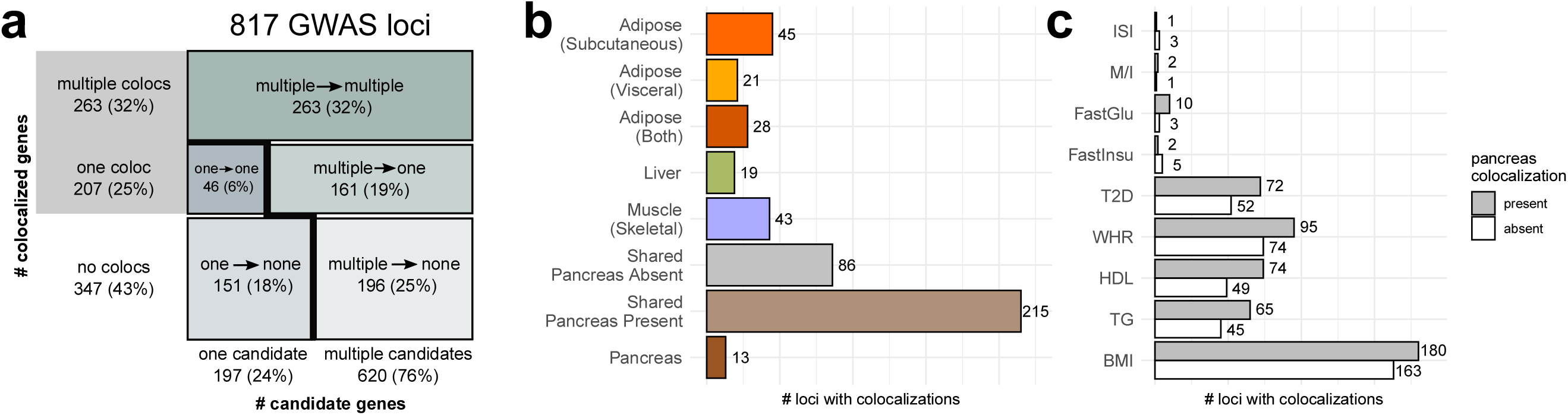
Colocalized loci show diverse causal architectures. (A) Mosaic plot categorizing all GWAS loci by the number of candidate genes (genes with GWAS signals overlapping QTLs) and the number of colocalized genes. Box sizes are proportional to the number of loci in the category. (B) Counts of tissue-specific and tissue-shared loci. (C) Total number of colocalized loci for each tested trait, separated by presence or absence of pancreas colocalization.

Of the 620 loci starting with multiple candidate genes, 26% showed just one colocalized gene, while 42% showed multiple colocalized genes, suggesting that GWAS loci might often harbor several causal genes. Indeed, we observed that some such loci contained multiple independent association signals that are located nearby on the genome but are neither LD-linked nor strictly overlapping. For example, a locus on chromosome 3 spanning 2.5 Mb contained not only a T2D-associated variant in an intron that colocalized with *RBM6*, but also a fasting glucose variant, located 500kb upstream of the *RBM6*-associated variant, regulating *MST1* expression and splicing (Fig. S5). At other loci, the various colocalized genes had QTL association signals that were both overlapping and LD-linked. For some of these loci, multiple co-regulated genes are functionally relevant to the disease, e.g., the *FADS1* / *FADS2* / *FADS3* locus [26] (Fig. S5). For others, one of the colocalized genes may be the driver of disease risk, while the other genes may be co-regulated with the causal gene but not directly relevant to the colocalized trait. For example, at the well-studied *SORT1* locus, functional experiments have proven a causal role for *SORT1* in regulating lipid levels but saw none for *PSRC1*, another LD-linked and colocalized gene [27] (Fig. S5). While these loci with multiple implicated genes are likely important contributors to disease risk, the ability of colocalization analyses to disentangle their roles is limited, and thus we subsequently focused on loci with only one colocalized gene.

### Tissue specificity dissects different components of disease

Previous work has used tissue specificity to inform tissues of action for causal genes [28], and others have further partitioned cardiometabolic risk loci into groups with primary roles in the pancreas, liver, adipose tissues, and others [6]. We hypothesized that genes colocalized exclusively in a single tissue might similarly form functional subgroups. Among loci with a single colocalized gene, we identified 30 subcutaneous adipose-specific (e.g. *LPL* and *PDGFC*; see Fig. 3A), 14 visceral adipose-specific (e.g. *NUP133* and *HPGDS*), 18 liver-specific (e.g. *SLC22A3* and *PNPLA3*), 19 skeletal muscle-specific (e.g. *CDKN2C* and *HMGB1*), and 5 pancreas-specific loci (e.g. *RYBP* and *CTRB2*). We found 16 additional (visceral and subcutaneous) adipose-specific loci including *PLEKHA1* (*TAPP1*), which is known to affect insulin sensitivity through its effect in adipose tissue [29] (Fig. 3A-B, Additional Data S4). Among tissue-specific colocalizations, the most muscle- and pancreas-specific colocalizations were associated with the glycemic traits T2D, fasting insulin, and fasting glucose; the most adipose-specific colocalizations with WHR; and the most liver-specific colocalizations with levels of HDL and TG (Table S5), in accordance with heritability enrichments for the same traits in a recent study [30].

**Figure 3:**
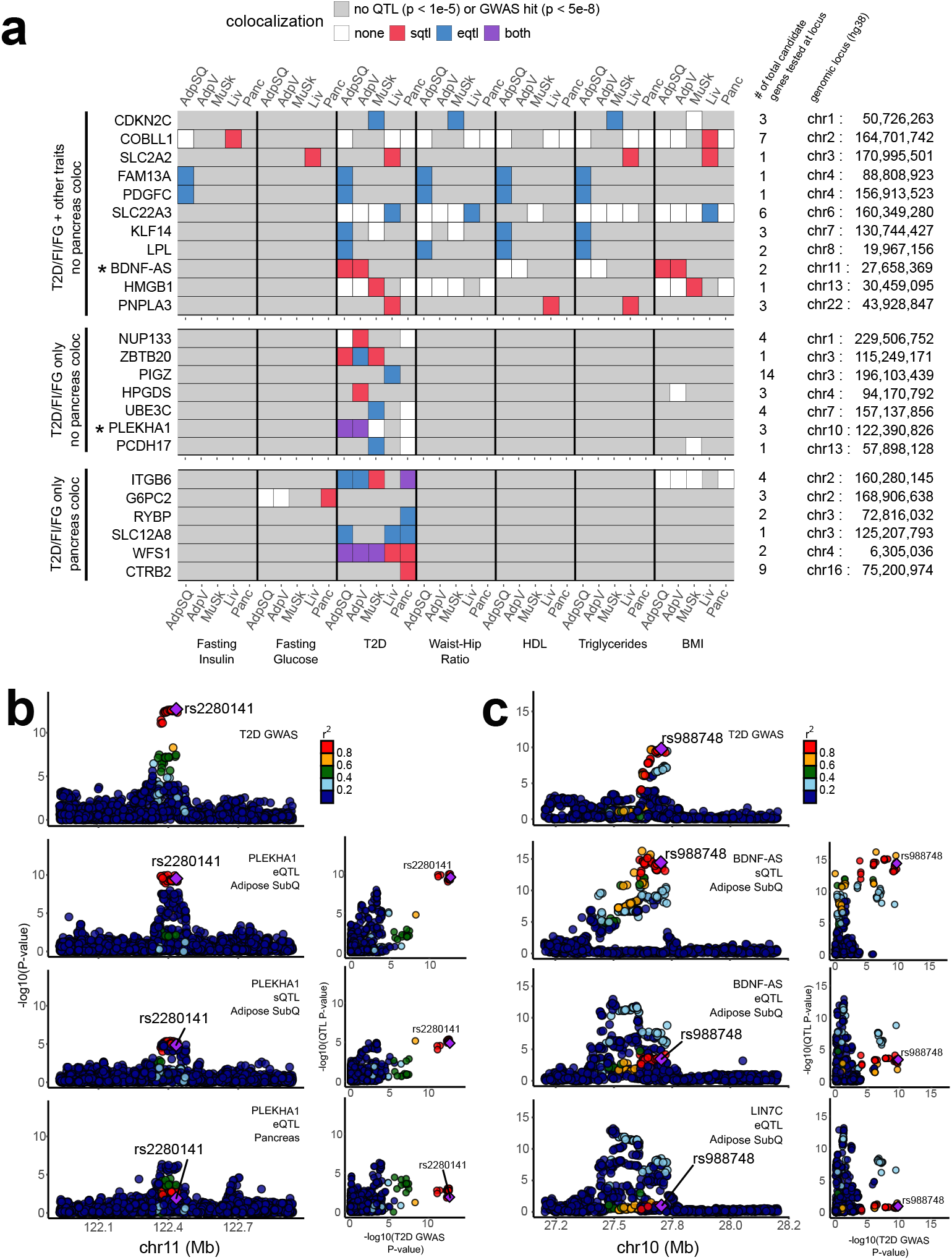
Post-colocalization follow-up identifies various patterns of shared colocalization across tissues and traits. (A) Unique genes colocalizing with T2D, fasting insulin, or fasting glucose GWAS at selected loci (each row represents one locus). (B) LocusCompare plots illustrating an adipose-specific colocalization in *PLEKHA1* (*TAPP1*). (C) LocusCompare plots illustrating an sQTL-specific colocalization for *BDNF-AS*. The eQTL association signal for another nearby gene, *LIN7C*, is shown for comparison.

To zoom in from the bulk tissue level to a finer resolution of the biological pathways and cell types in which colocalized genes are active, we tested these genes for overlap with cell type-specific genes we inferred from the Human Cell Landscape [31] using pSI [32, 33] and for membership in co-expression modules that we generated from GTEx using weighted gene co-expression network analysis [34, 35] (Additional Data S5 & S6, see Methods). For example, the *LPL* (lipoprotein lipase) gene, whose eQTLs colocalize with fasting insulin, WHR, HDL, and TG exclusively in subcutaneous adipose tissue in GTEx, was identified as a cell type-specific gene for adipocytes, in contrast with other adipose-colocalized genes that were specific to mast cells (e.g. *HPGDS*) and neutrophils (e.g. *EPC2*). Furthermore, *LPL* is a member of a GTEx co-expression module associated primarily with fatty acid metabolism and biosynthesis pathways, in accordance with the gene’s known functions [36]. As another example, the *FGFR1* gene colocalized with T2D exclusively in GTEx muscle tissue and was ascribed to fibroblasts, myogenic precursor cells, and natural killer (NK) cells. In the GTEx co-expression networks, *FGFR1* belongs to a module associated with extracellular matrix organization, collagen formation, and cell adhesion.

Tissue specificity can further dissect different components of disease. Cardiometabolic functions associated to IR are more likely to be mediated by loci colocalized only in non-pancreas tissues, while loci colocalizing with pancreas may act through molecular mechanisms related to insulin production or secretion. Tissue-specific loci outside of the pancreas comprised a third of all colocalized loci (156 of 470, 33%), implicating a plethora of potential IR candidate genes. Even among the 301 loci with tissue-shared colocalization, 86 had no colocalization detected in pancreas (Fig. 2B), such as the locus associated with *ZBTB20* (Fig. 3A), further increasing the number of potential IR candidate genes.

### Sharing across traits places causal genes within functional disease subgroups

Previous joint analyses of T2D along with similar traits have sorted GWAS loci and coding variants into subgroups [5, 6, 7, 8, 9] representing different components of cardiometabolic disease biology. These subgroups include insulin production- and secretion-related clusters deemed “proinsulin” and “*β*-cell” clusters, as well as “obesity”, “lipodystrophy”, and “liver/lipid” clusters [6]. Thus, we used our colocalization results to distinguish between candidate causal genes belonging to specific subgroups. Among loci colocalized in pancreas, we observed several genes assigned previously to the *β*-cell cluster, indicating a likely role in dysfunctional insulin production or secretion [6]. *CTRB2*, for example, colocalized with T2D exclusively in pancreas, giving further credence to its previous placement in this cluster [6] (Fig. 3A). Similarly, other genes with pancreas-specific colocalization, such as *RYBP* and *G6PC2*, may also contribute to the *β*-cell cluster.

Sharing across traits also informs the functional subgroup of a colocalized gene. For example, we saw adipose-specific TG and HDL colocalizations for both *KLF14* and *LPL* (Fig. 3A), two genes assigned previously to a lipodystrophy cluster [6] that has been shown to overlap with IR biology. *LPL* further colocalized with WHR in adipose tissue, the main tissue characterizing the lipodystrophy phenotype. By contrast, we found liver-specific colocalizations with T2D, TG, and HDL in *PNPLA3*, which was previously assigned to a cluster involving liver/lipids and lower overall TG levels [6]. Thus, T2D-colocalized genes that also colocalized in non-pancreas tissues with TG/HDL or with WHR are likely candidates for either the liver/lipids or lipodystrophy clusters, respectively. Furthermore, tissue specificity can reinforce the trait sharing-based categorization. A gene such as *PDGFC* that colocalizes with T2D, fasting insulin, WHR, HDL, and TG, all in adipose tissue, is a stronger candidate for a role in lipodystrophy, whereas a gene like *SLC2A2* with liver colocalization for T2D, fasting glucose, triglycerides, and BMI may be more relevant to the liver/lipids cluster.

Among all loci containing a single colocalized gene, 13 colocalized with more than one broad trait category (glycemic, anthropometric, or lipid traits) (Fig. 4A). These loci harbor several previously known cardiometabolic causal genes such as *KLF14* and *LPL*, but also novel candidates such as *PDGFC*. The relative directional effects of these genes across traits reflected known relationships between traits; i.e., genes whose expression was associated with higher risk and/or levels of IR, T2D, WHR, fasting insulin, fasting glucose, and TGs were generally also associated lower levels of HDL, and vice-versa (Fig. 4B, Fig. S6, & Additional Data S7). Similarly, they confirmed the directions of several previously studied genes; for example, decreased expression of *KLF14* correlated with increased risk of T2D, which has also been demonstrated previously in subcutaneous adipose tissue [37]. Moreover, these genes’ directions of effect are important for drug development, as they indicate whether inhibition or activation of a genetic target will be therapeutically beneficial.

**Figure 4:**
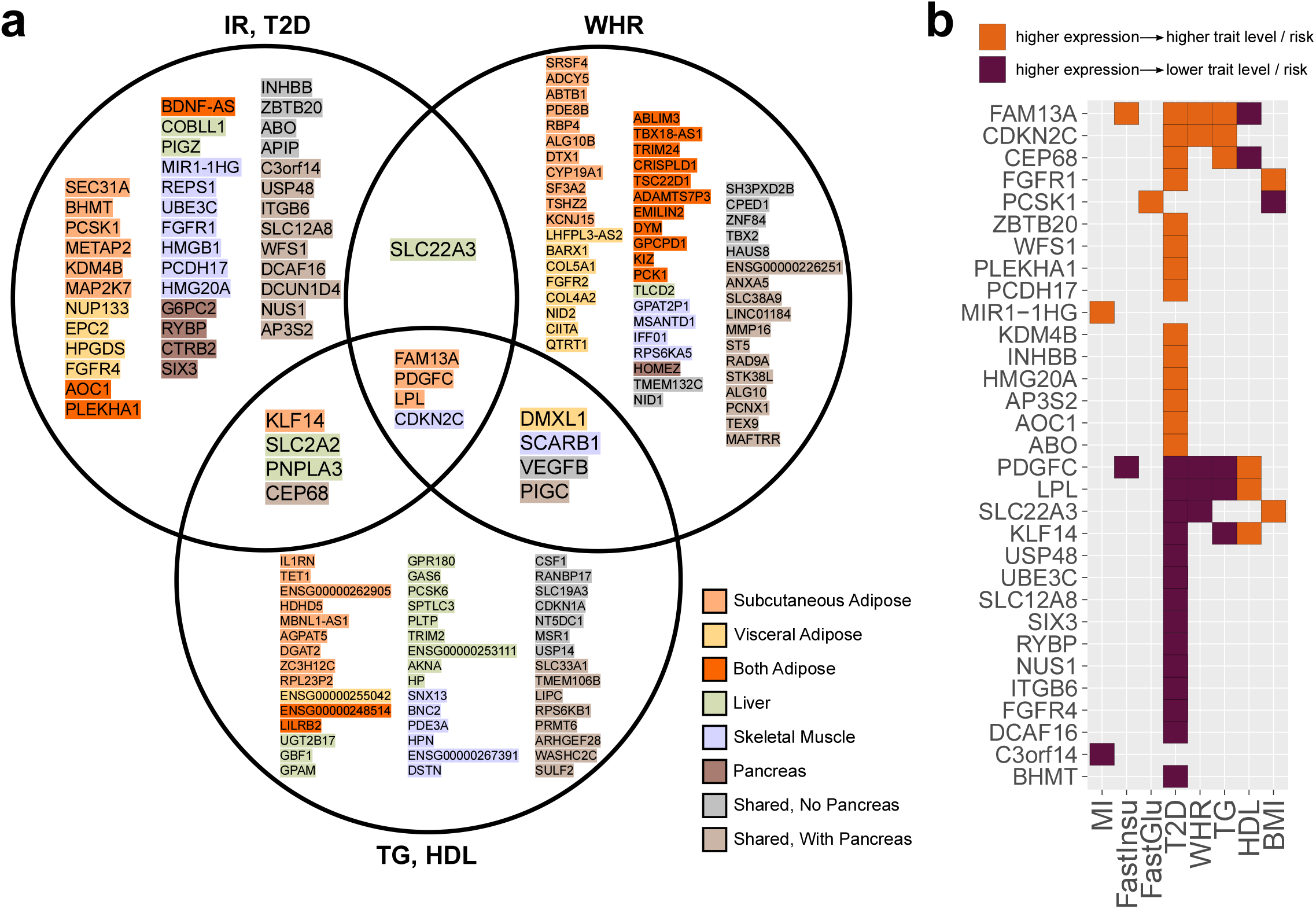
A prioritized set of uniquely colocalized candidate genes for follow-up testing. (A) The set of genes that are uniquely colocalized at their respective loci with insulin sensitivity, fasting glucose, fasting insulin, T2D, WHR, TG, or HDL. The highlight color indicates the tissues of colocalization. (B) Directions of effect of uniquely colocalized IR / T2D genes on GWAS trait risks and levels.

### Perturbation with physiological and pharmacological regulators contextualizes candidate causal genes

Beyond identifying causal genes, it is critical to place them within a broader systemic model of disease mechanisms. To this end, we tested every candidate causal gene for differential expression under 21 physiological and pharmacological cardiometabolic regulators in human adipocytes, hepatocytes, and skeletal muscle cells [30] (Table S6). We thus generated a canvas of upstream molecular signals controlling the expression of candidate causal genes in relevant metabolic contexts (Fig. 5A, Fig. S7, & Additional Data S8). Of the 152 uniquely colocalized genes for IR/T2D, WHR, and TG/HDL, 42 were regulated by insulin and 35 by glucose, including 17 regulated by both, pointing to clear upstream regulators in the context of disease. Other metabolic perturbations regulated expression of 30 more genes not regulated by glucose or insulin. For example, the uniquely colocalized fibroblast growth factor receptors *FGFR1* and *FGFR4* both show increased expression in response to insulin-like growth factor 1 (IGF1) in skeletal muscle cells, while they exhibit decreased expression in human hepatocytes in response to the glucocorticoid dexamethasone, which inhibits insulin signaling in other systems [38, 39].

**Figure 5:**
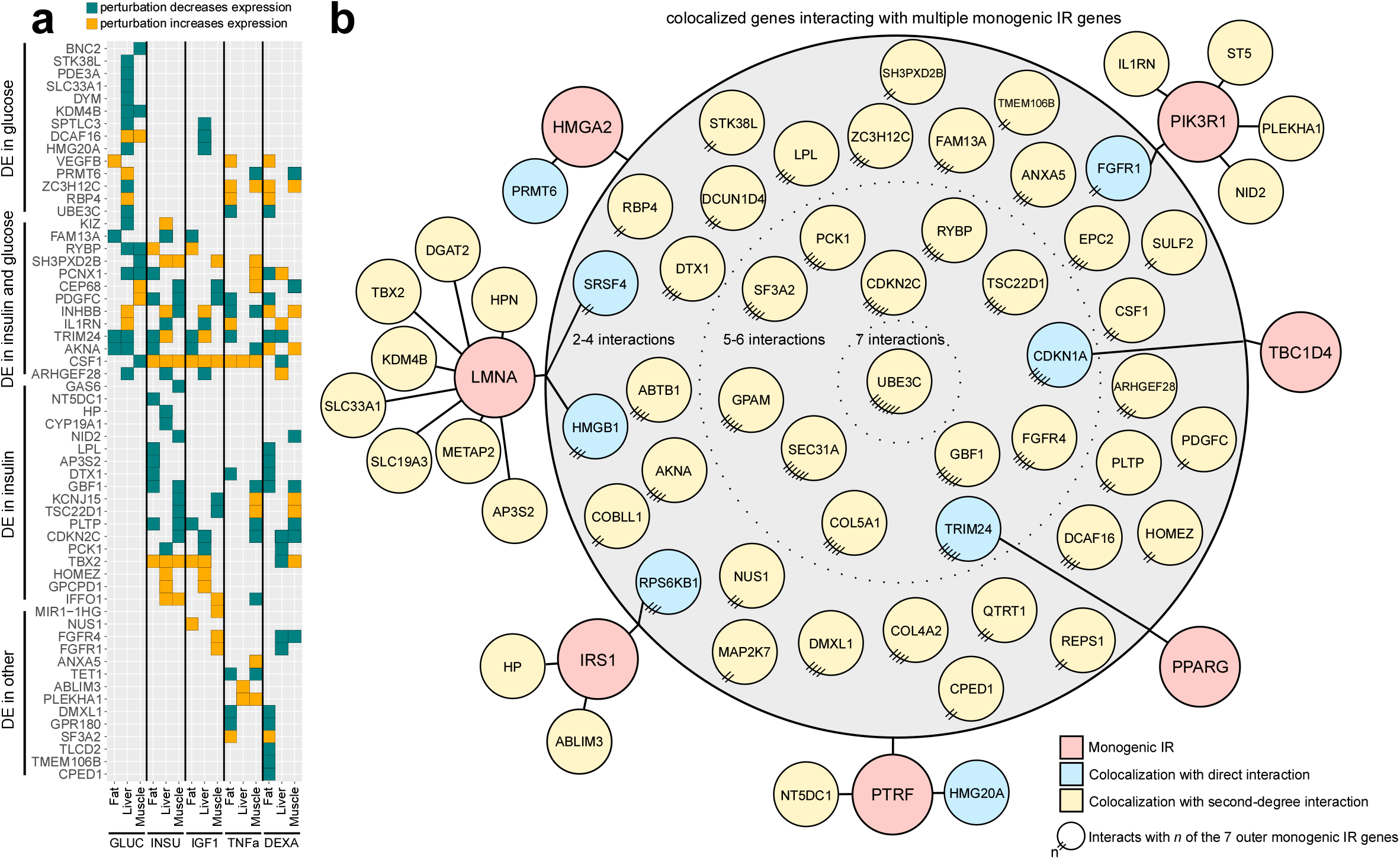
Genes prioritized through colocalization analysis respond to metabolic perturbations. (A) Candidate causal genes with altered expression under five metabolic perturbations (FDR *<* 5%). (B) PPI network interactions in perturbation conditions between uniquely colocalized genes and seven known monogenic IR or T2D genes. Colocalized genes in blue nodes interact directly with the monogenic IR/T2D genes in red nodes; colocalized genes in yellow nodes interact via one intermediary gene. All genes in the central gray circle interact with multiple monogenic IR/T2D genes on the periphery, and genes closer to the center interact with more of the monogenic IR/T2D genes. To simplify visualization, not all direct links are depicted for genes with multiple interactions.

Effects of causal genes can be further modified by pharmacological intervention. For 30 of our candidate causal genes, we observed a response to atorvastatin, metformin, or rosiglitazone, three drugs used for the treatment of cardiometabolic diseases. For instance, *COBLL1*, a gene with liver-specific colocalization for fasting insulin and BMI and previously associated with non-alcoholic fatty liver disease [40, 41], showed decreased expression under both atorvastatin and metformin in liver. Another gene, *GPAM*, which colocalized with HDL and TG also in liver, showed reduced expression under all three of these treatments (atorvastatin and metformin in liver, rosiglitazone in fat cells).

We hypothesized that our uniquely colocalized genes might also interact with other key cardiometabolic genes regulated under the same conditions. Starting from the full list of known protein-protein interactions (PPIs) identified in BioGrid [42], we pruned this list to define perturbation-specific PPI networks of protein-coding genes active (differentially expressed) under each perturbation (Additional Data S9). We then identified interactions between candidate causal genes and a curated list of 49 known IR or T2D gene(s) (Tables S7, S8). The resulting network of known and candidate IR/T2D genes (Fig. 5B, Fig. S8 & Additional Data S10) revealed a tight web of connections between our colocalized genes and those known IR/T2D genes. Eight candidate causal genes interact directly with an IR/T2D gene (7 total) in at least one perturbation condition, and 54 other candidates interact via one intermediary protein (Fig. 5B). For example, the fibroblast growth factor receptor *FGFR1* interacts directly with the known monogenic IR kinase *PIK3R1* [43], and its family member *FGFR4* also interacts with several IR/T2D genes (Fig. 5B). Together, these results showcase the value of studying colocalization-based candidate causal genes within the appropriate cellular contexts, and in our case, under metabolically relevant cell-extrinsic signals as part of a broader network of disease associated genes.

### High-priority list of causal candidate genes for cardiometabolic disease

To facilitate informed selection of candidate causal genes for follow-up, we summarize in Fig. 6 and Fig. S9 our complete list of uniquely colocalized genes for IR/T2D, WHR, and TG/HDL, which represent both tissue-specific and tissue-shared candidate causal genes. These associations can be exclusive or overlapping among eQTLs and sQTLs, and in some instances shared across metabolic traits. Moreover, we provide directions of effect on metabolic traits, empirical data on potential upstream regulators, and mechanistic insights through PPI networks.

**Figure 6:**
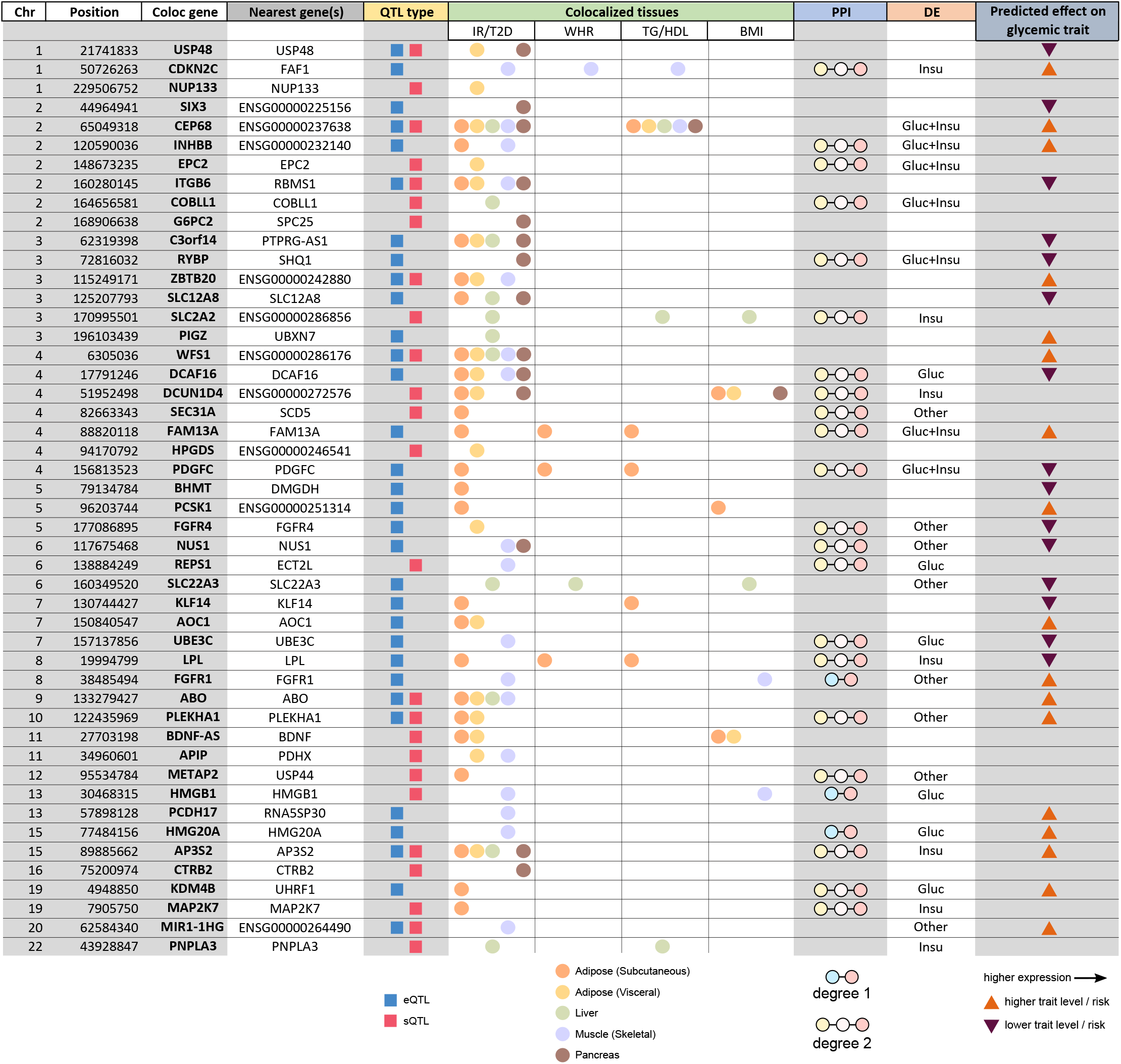
Integrative summary of causal evidence at 48 IR / T2D loci with a single colocalized gene. All loci with a colocalization for type 2 diabetes, fasting glucose, fasting insulin, and/or insulin sensitivity are included.

Using these results, we can prioritize candidate causal genes, further dissecting the set based on additional relevant features. As an example, the previously characterized cardiometabolic risk gene *FAM13A* [17] not only colocalizes in subcutaneous adipose tissue with fasting insulin, T2D, WHR, TG, and HDL, but it also has second-order PPI interactions with 10 known IR/T2D genes and is differentially expressed under both glucose and insulin stimuli. Similarly, another gene, *PDGFC*, which has been studied in the context of angiogenesis [44] and vascular diseases [45] but not yet extensively within IR/T2D, colocalizes in the same tissue (subcutaneous adipose) with all the same traits as *FAM13A*, although its direction of effect is opposite to *FAM13A*. *PDGFC* also responds to glucose and insulin stimuli and interacts with six known IR/T2D genes, two of which also interact with *FAM13A* (*CAV1* and *ZMPSTE24*). As one more example, we highlight the gene *CDKN2C*, which colocalizes in muscle with T2D, WHR, and TG despite that the GWAS SNP lies directly within an intron of another gene, *FAF1*. *CDKN2C* has seven PPI interactions with known IR/T2D genes and is differentially expressed under glucose stimulation. These and other examples demonstrate that the combination of our newly implemented colocalization approach with our large-scale perturbation data in metabolic cell types provides a testable list of highly probable causal genes for GWAS loci in the context of IR, T2D, and the associated cardiometabolic traits.

## Discussion

Cardiometabolic diseases have now reached staggering levels around the globe. After almost two decades of GWAS and the discovery of hundreds of loci associated with cardiometabolic traits, few causal genes have been described in the context of IR, which is a key underlying condition of cardiometabolic disease. Our approach is the first of its kind to integrate colocalizations across traits, tissues, and QTL types with experimental perturbations to prioritize candidate genes for human disease. Though the GWAS traits and QTL tissues used here are specifically relevant to cardiometabolic disease, this approach is broadly applicable to many other human complex diseases and traits.

Our colocalization analysis incorporating both eQTLs and sQTLs found single colocalizations for 19% of the loci and multiple colocalizations in 25% of the loci analyzed. Teasing apart the contribution of individual genes to loci with multiple colocalizations remains an additional challenge. However, at loci with a single colocalization, we identify 152 candidate causal genes for IR/T2D, WHR, and TG/HDL. Our list is supported by colocalization of genes known to lead to Mendelian forms of diabetes (e.g., *SLC2A2* and *WFS1*) [46, 47] and of genes with previously demonstrated mechanisms of association with IR/T2D (e.g. *METAP2*, *PLEKHA1* [*TAPP1*], *FAM13A*, and *KLF14*) [48, 29, 49, 37]. Our approach also points at novel, shared genetic architecture of traits. For example, two genes previously associated with non-alcoholic fatty liver disease, or NAFLD (*COBLL1* and *PNPLA3*) [40, 41], show colocalization in liver (Fig. 6), the former with fasting insulin and BMI and the latter with T2D, HDL, and TG, implicating them as shared genetic associations for IR/T2D and NAFLD. Similarly, *BDNF-AS* colocalizes with T2D and BMI in adipose tissue and may link obesity, adiposity and mood disorders [50, 51]. Finally, the FGFR family contributes broadly, with *FGFR1* colocalized in skeletal muscle for IR/T2D, *FGFR4* in visceral adipose tissue for IR/T2D, and *FGFR2* in visceral adipose tissue for WHR, highlighting the relevance of this family of receptors in metabolic regulation [52]. Based on our approach, the different patterns of shared colocalizations across tissues and traits suggest how these genes fit into the broader landscape of human complex disease, and in the context of cardiometabolic disease, categorize the novel candidate causal genes into potential IR- and non-IR-related subgroups.

We identified upstream extrinsic regulators of the candidate causal genes through a large-scale gene expression assay of metabolically relevant signals, and using PPI networks we further connected these candidate genes to the broader network of other previously established IR and T2D genes. The resulting regulators and interactors for each candidate causal gene are a starting point to investigate the molecular mechanisms linking these genes to cardiometabolic disease. Moreover, it is enticing to consider the different mechanistic implications for the subgroups of colocalized genes regulated by different perturbations. On one hand, candidate causal genes regulated by insulin and/or glucose may be an integral part of the core metabolic signaling and transcriptional network associated to insulin sensitivity, glucose homeostasis and cardiometabolic trait regulation. On the other hand, those candidate genes regulated by any other perturbation (including pharmacological regulators) may reflect parallel signaling and transcriptional networks with a significant regulatory role or crosstalk with the core insulin/glucose network, as with for example the FGFR family members *FGFR1*, *FGFR2* and *FGFR4*. By contextualizing candidate causal genes, our perturbation analysis strengthens the interpretation of the colocalization results that bridge the gap from GWAS variants to actionable causal genes.

## Conclusions

Our integrative list of high-confidence cardiometabolic genes is both a general resource for investigators and a tool for detailed dissection of genes into relevant disease subgroups. Together, the integration of these multi-tissue and multi-trait colocalization results with their upstream extrinsic regulators provides extensive, contextual gene-by-gene annotations for genes involved in IR, T2D, and associated cardiometabolic traits and will enhance drug development for cardiometabolic diseases.

## Methods

### Preprocessing of GWAS and QTL files

We downloaded publicly available association summary statistics for 9 cardiometabolic traits (12 total GWAS, Table S2) and GTEx v8 eQTLs and sQTLs summary statistics for five relevant human tissues, i.e. adipose visceral and subcutaneous, skeletal muscle, liver, and pancreas (Table S1). Unless otherwise specified, eQTLs and sQTLs were analyzed identically in all subsequent steps, except that the feature of interest for sQTLs is the number of splice events detected at a single intragenic splice junction rather than the number of transcripts detected for a single gene. All splice junctions were already assigned to a single gene (Ensembl ID) in the GTEx v8 data.

We used the *gwas-download* toolkit https://www.github.com/mikegloudemans/gwas-download to sort, consistently re-format, and generate *tabix* index files for each of the GWAS and QTL summary statistics files.

### Selection of overlapping GWAS and QTL loci for colocalization tests

Here we define a “colocalization test locus” as a unique combination of a specific GWAS trait, a locus of the genome, a QTL tissue, and a gene measured within that tissue. The goal of the colocalization test is to determine whether the GWAS signal matches the QTL signal for that gene in that tissue: that is, whether the GWAS trait and gene expression share a common genetic causal variant. Because the total number of loci and QTL genes in the genome is large and therefore computationally expensive to test, and to minimize the potential for false positives, we limited our analysis to loci that have both a GWAS association and a QTL association in a specified tissue and gene. For two loci to be considered independently, we required them to be located at least 500kb apart. To increase our sensitivity to relevant loci, we set these thresholds at GWAS P *<* 5e-8 and QTL P *<* 1e-5. For two traits directly measuring insulin response (M/I and insulin sensitivity index), which were limited in GWAS power by small sample sizes, we lowered the GWAS significance threshold to P *<* 1e-5 to increase sensitivity, since any spurious GWAS loci are unlikely also to pass the subsequent colocalization tests. This selection process is depicted in Fig. S1 and is also implemented in the ‘gwas-download’ GitHub repository linked above in the section “Preprocessing of GWAS and QTL files.”

### Clustering of GWAS lead SNPs into individual loci with LDetect

To determine which nearby GWAS signals for different traits were part of the same genomic locus, we partitioned the genome into 1,724 loci. We used a pre-defined set of LD-independent regions (LDetect, using European-derived LD regions, Additional Data S2) [24]; an advantage to this approach is that the resulting loci are invariant to the number of traits and tissues included in the colocalization analysis. We note that it is possible for a single locus as defined by LDetect still to contain multiple GWAS associations for the same GWAS trait, as long as they are 500kb apart.

### Colocalization testing

Colocalization analysis computes the probability that genetic association signals for a GWAS trait and a QTL feature are produced by a common causal variant, and importantly removes misleading signals with incidental GWAS-QTL overlaps due to complicated LD tagging patterns [53]. Several methods have been designed for this purpose, such as eCAVIAR [54], which produces an intuitive colocalization posterior probability (CLPP) score, directly indicating the probability of a shared causal variant between a tested GWAS and QTL study. One limitation we observed with this approach, however, is that it becomes overly conservative when several assayed variants are in near-perfect LD with the true causal variant, in which case it yields very low probabilities even for loci where the causal gene is known (e.g. *WFS1*, see Fig. S3).

To address these limitations, we performed our colocalization analysis using a novel custom integration of the FINEMAP [55] and eCAVIAR [54] methods. For each previously selected test case (see “Selection of overlapping GWAS and QTL loci for colocalization tests”), we narrowed our summary statistics to the set of the SNPs tested for association with both the given GWAS trait and the given QTL trait, and removed all sites containing less than 10 SNPs after this filter. Using the full 1000 Genomes dataset from phase 3 (2,504 individuals) as a reference population [56], we estimated LD between every pair of SNPs. We then ran FINEMAP [55] independently on the GWAS and the QTL summary statistics to obtain posterior probabilities of causality for each of the remaining SNPs, and we combined these probabilities to compute a colocalization posterior probability (CLPP) using the formula described in the eCAVIAR method [54].

Because the canonical CLPP score is highly conservative in regions with densely profiled, high-LD SNPs, we modified the score formula to produce an LD-modified CLPP score, which we refer to as the *CLPP_mod_* score.

The original CLPP is defined as

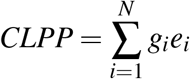

Where

- *g_i_* is the probability that the ith SNP is the causal variant for the GWAS trait
- *e_i_* is the probability that the ith SNP is the causal variant for the QTL trait
- *N* is the total number of variants at the locus.

Our LD-modified CLPP score is a generalization of this score, given by

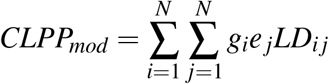

Where *LD_i j_* is the LD (*r*^2^) between the *i*th and the *j*th SNP in a reference population.

This modified approach produces an LD-modified CLPP (colocalization posterior probability) score, the *CLPP_mod_* score. It intuitively represents the sum over joint causal probabilities across all pairs of GWAS + QTL SNPs at the locus, with each pair’s contribution to the final score weighted by the LD between these two SNPs. Like the original *CLPP* score, the *CLPP_mod_* at a locus will always be between 0 and 1. Subsequent visual inspection of juxtaposed GWAS and QTL Locus-Compare plots at high and low *CLPP_mod_* score loci confirmed that our LD-modified CLPP score detects true colocalized loci, but without disproportionately penalizing high-LD loci (Fig. S3).

### Quantification of number of genes and loci tested / colocalized

We counted the total number of GWAS loci and expressed genes (protein-coding and others) selected for each locus before filtering to the genes with overlapping QTLs. We additionally determined the number of independent loci across all included GWAS traits by grouping nearby loci for different traits into the same numbered locus with LDetect, as described above [24]. Given that a typical locus has 20-50 genes located within 1 Mb, the number of candidate genes is quite large. We then quantified the effect of filtering locus-gene pairs to those in which the lead GWAS SNP is a significant eQTL or sQTL for that gene in at least one of our five QTL tissues (p *<* 1e-5). We recomputed the number of loci and genes for the filtered set. Finally, we computed the number of loci and genes one more time for those loci and genes colocalizing with at least one trait in one tissue (*CLPP_mod_ >* 0.35, representing the top 20% of all tested combinations). The numbers of genes and loci passing each of these filtering steps are shown in Fig. 1A and Fig. S2. When summarizing the colocalization results in heatmaps for Fig. 3, the results from all four of the T2D studies were collapsed into a single column representing the top colocalization score in any study.

### Definition of tissue specificity

A gene was considered tissue-specific if had colocalizations in one tissue, but colocalized in no other tissues for any trait. A locus was considered tissue-specific if it contained a gene or genes with colocalizations in one specific tissue, but no genes colocalized in any other tissues for any trait.

### GTEx coexpression modules and cell type specificity

GTEx coexpression modules are described in a previous publication [35]. We treated the genes in each module as a gene set, and tested for functional enrichment using clusterprofiler [57] with candidate enrichments from consensusDB [58]. (See Additional Data S6 for a list of all modules and enrichments.)

### Assigning directional effects to colocalized loci

For all loci with a single colocalized eQTL gene, we compared the effect direction of the lead variant on expression with its effect direction on the risk or level of the colocalized GWAS trait. Some alignment was required to ensure consistency of the reported effect alleles between the QTL and GWAS summary statistics files. For each colocalization, we thus determined whether an increase in the eQTL target gene’s expression was associated with an increase or decrease in the GWAS trait risk or level, as reported in Fig. 4B. The full list of all directional alignments for uniquely colocalized eQTL genes is given in Additional Data S7. This analysis applies only to eQTL colocalizations since sQTLs do not have a naturally interpretable direction of increased or decreased expression.

### Perturbation experiments

We tested our list of uniquely colocalized genes for differential expression under 21 metabolic perturbations in human adipocytes (SGBS), hepatocytes (HepG2), and skeletal muscle cells (HMCL-7304). The experimental design has been previously described and applied to prioritize how these perturbations impact diverse complex traits [30]. All perturbations are listed in Table S6, and the intersection of these data with uniquely colocalized cardiometabolic genes is shown in Fig. 5A, Fig. S7, & Additional Data S8.

### Protein-protein interaction networks

We obtained a list of experimentally confirmed protein-protein interactions from the BioGRID public database [42]. For each of the 63 IR-relevant perturbations, we constructed a pruned PPI network with *igraph* [59] consisting of only protein pairs that 1) interacted in the original PPI network, and 2) were both differentially expressed under the given perturbation condition, indicating that they likely interact within that context.

Once these pruned PPI networks were obtained for each perturbation context, we determined all uniquely colocalized genes that interact in these networks with one or more of the previously reported monogenic IR/T2D genes or known T2D genes from genetic studies listed in Tables S7 and S8, either directly or via a single intermediary protein. We found 7 of these genes directly interacting with one or more of our uniquely colocalized genes, and in Fig. 5B, we depict all uniquely colocalized genes demonstrating first- and second-order interactions with any of these 7 IR/T2D genes. An additional 22 monogenic IR/T2D genes had exclusively second-order or higher interactions with uniquely colocalized genes; these genes are not depicted in the figure for readability. A complete list of these and all higher-order interactions is given in Additional Data S9.

## Data Availability

The code for colocalization analysis is given at https://www.github.com/mikegloudemans/insulin-resistance-colocalization. The RNA-seq data for perturbation experiments have been uploaded to GEO with accession number GSE179347. Colocalization heatmaps depicting all genes tested at every locus in the style of Fig. 3A are publicly downloadable as PDFs from https://zenodo.org/record/4659095. Other results files are included as Additional Data Files, as referenced in the Results section. A pipeline for running our adaptation of the FINEMAP / eCAVIAR pipeline is at https://bitbucket.org/mgloud/production_coloc_pipeline, and a generalized toolkit for generating heatmaps for any sets of GWAS and QTL summary statistics is at https://github.com/mikegloudemans/post-coloc-toolkit.

https://bitbucket.org/mgloud/production_coloc_pipeline

https://github.com/mikegloudemans/post-coloc-toolkit/blob/master/README.md

https://zenodo.org/record/4659095

## Abbreviations

The following abbreviations are used in the text:

BMI: Body mass index
eQTL: Expression quantitative trait locus
FG: Fasting glucose
FI: Fasting insulin
GTEx: Genotype-Tissue Expression Consortium
GWAS: Genome-wide association study
HDL: High-density lipoprotein
IR: Insulin resistance
IS: Insulin sensitivity
LD: Linkage disequilibrium
NAFLD: Non-alcoholic fatty liver disease
PPI: Protein-protein interaction
QTL: Quantitative trait locus
SNP: Single-nucleotide polymorphism
sQTL: Splicing quantitative trait locus
T2D: Type 2 diabetes
TG: Triglycerides
WHR: Waist-hip ratio

## Declarations

### Ethics approval and consent to participate

Not applicable for this study.

### Consent for publication

Not applicable for this study.

### Availability of data and materials

The code for colocalization analysis is given at https://www.github.com/mikegloudemans/insulin-resistance-colocalization. The RNA-seq data for perturbation experiments have been uploaded to GEO with accession number GSE179347. Colocalization heatmaps depicting all genes tested at every locus in the style of Fig. 3A are downloadable as PDFs from https://zenodo.org/record/4659095. Other results files are included as Additional Data Files, as referenced in the Results section. A pipeline for running our adaptation of the FINEMAP / eCAVIAR pipeline is at https://bitbucket.org/mgloud/production coloc pipeline, and a generalized Snakemake toolkit for generating heatmaps for any sets of GWAS and QTL summary statistics is at https://github.com/mikegloudemans/post-coloc-toolkit.

### Authors’ contributions

MJG, BB, DN, EI, TQ, SBM, JWK, and ICO were involved in the conception and design of the work. MJG, BB, DN, and MGD performed statistical analyses. MJG, BB, and ICO were involved in acquisition and quality control of additional data. ICO reviewed and edited the manuscript, supervised the project and performed the perturbation experiments. MW provided the SGBS cells and methods for SGBS cell culture. MJG drafted the manuscript with input from BB, ICO, SBM and JWK. All authors contributed to the interpretation of results and critically reviewed the manuscript.

## Acknowledgments

We thank Pagé Goddard for sharing her design for Fig. S1.

## Supplementary Figures and Tables

**Figure S1:**
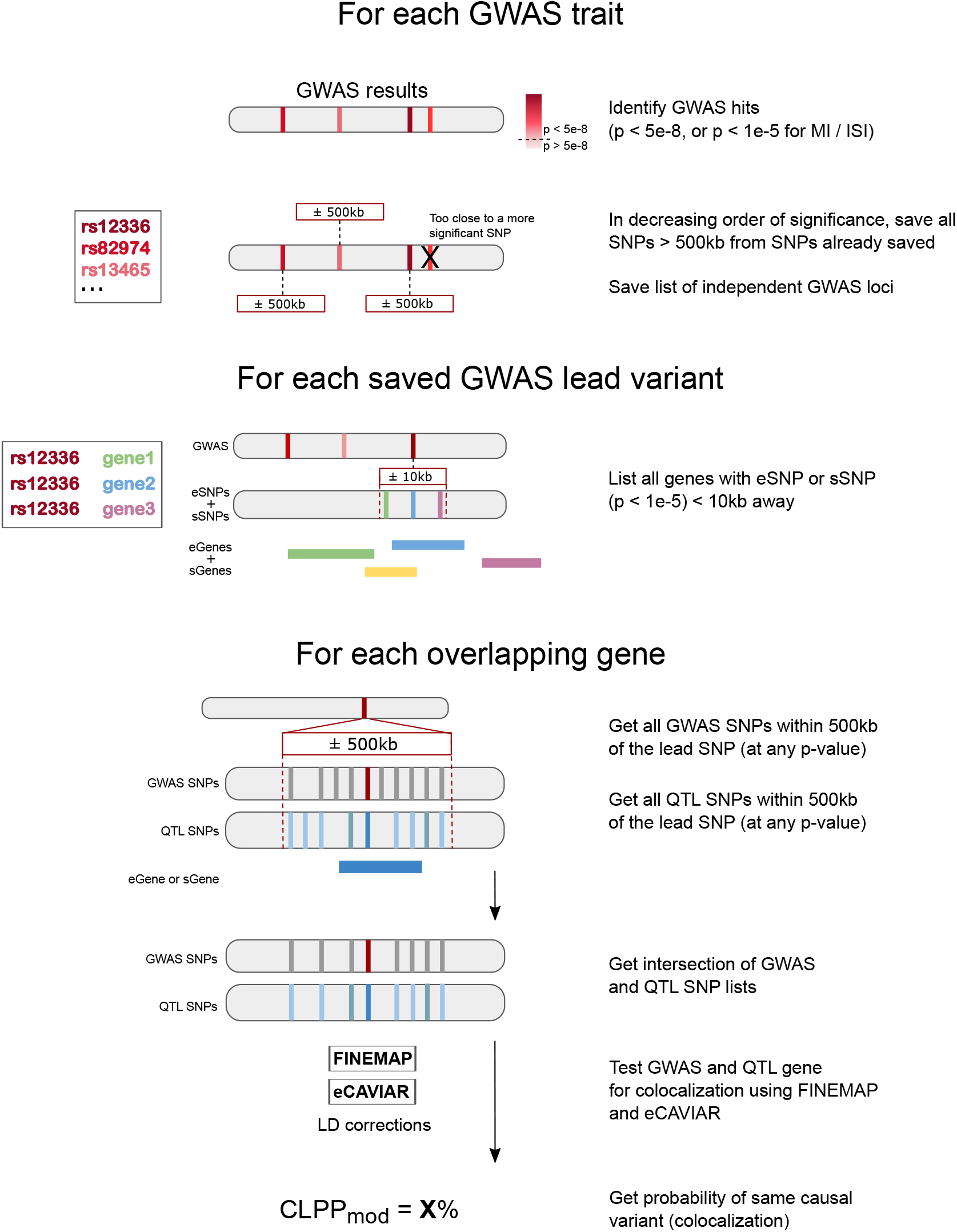
Selection of genome-wide significant loci and overlapping eQTL/sQTL features for colocalization testing.

**Figure S2:**
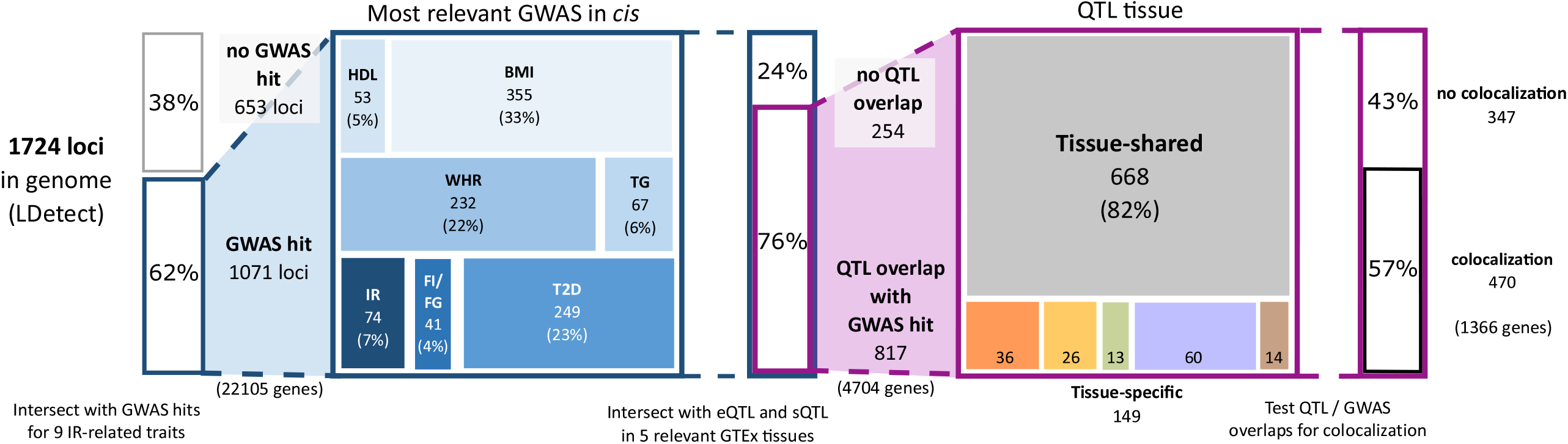
Number of loci per pre-colocalization step. We started with 1,724 genomic loci, encompassing the entire genome as segmented using the LDetect algorithm. 1,071 of these loci harbored a GWAS association for at least one of the cardiometabolic traits, and colocalization analysis identified at least one candidate causal gene at 470 of these loci.

**Figure S3:**
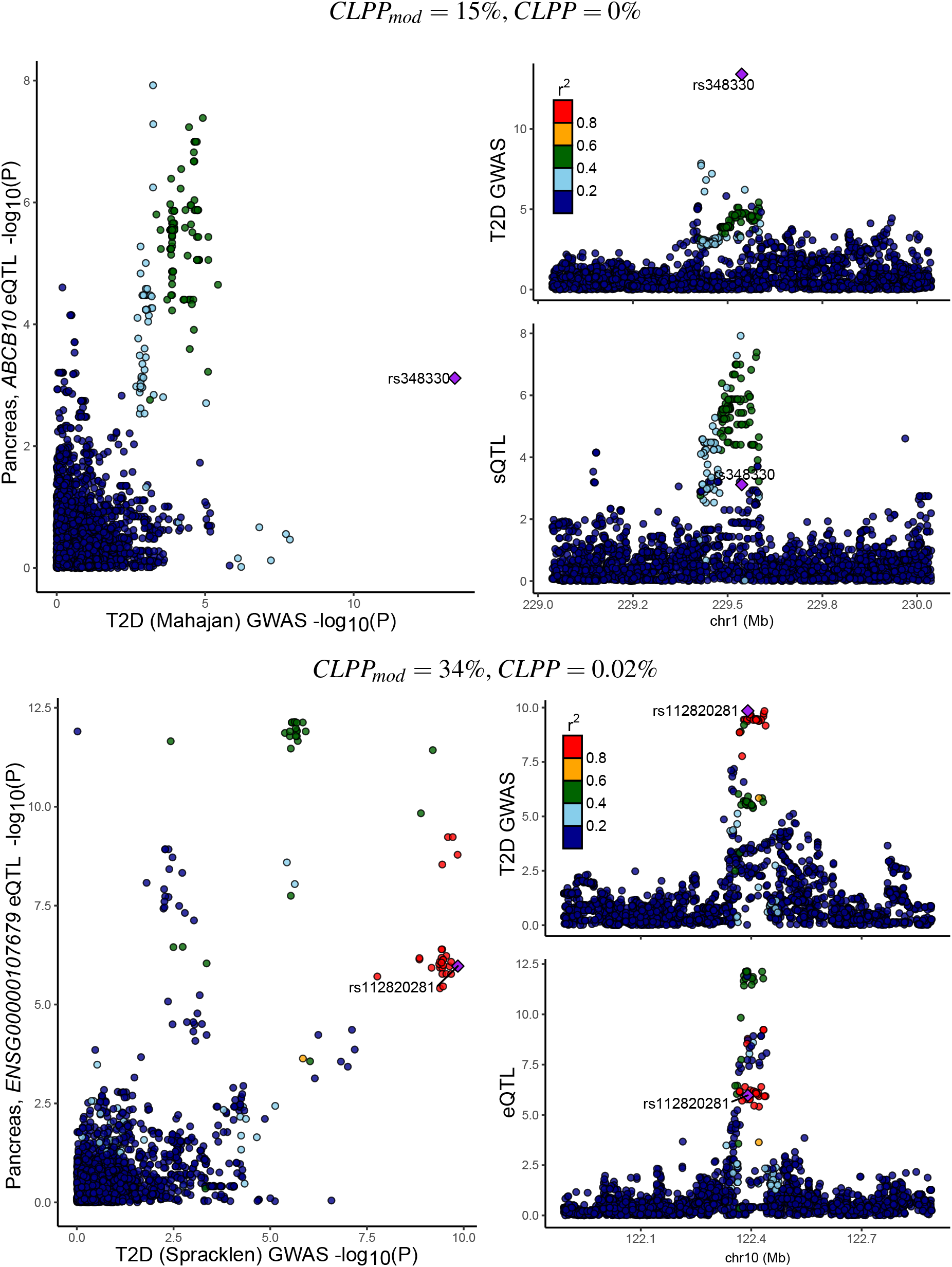
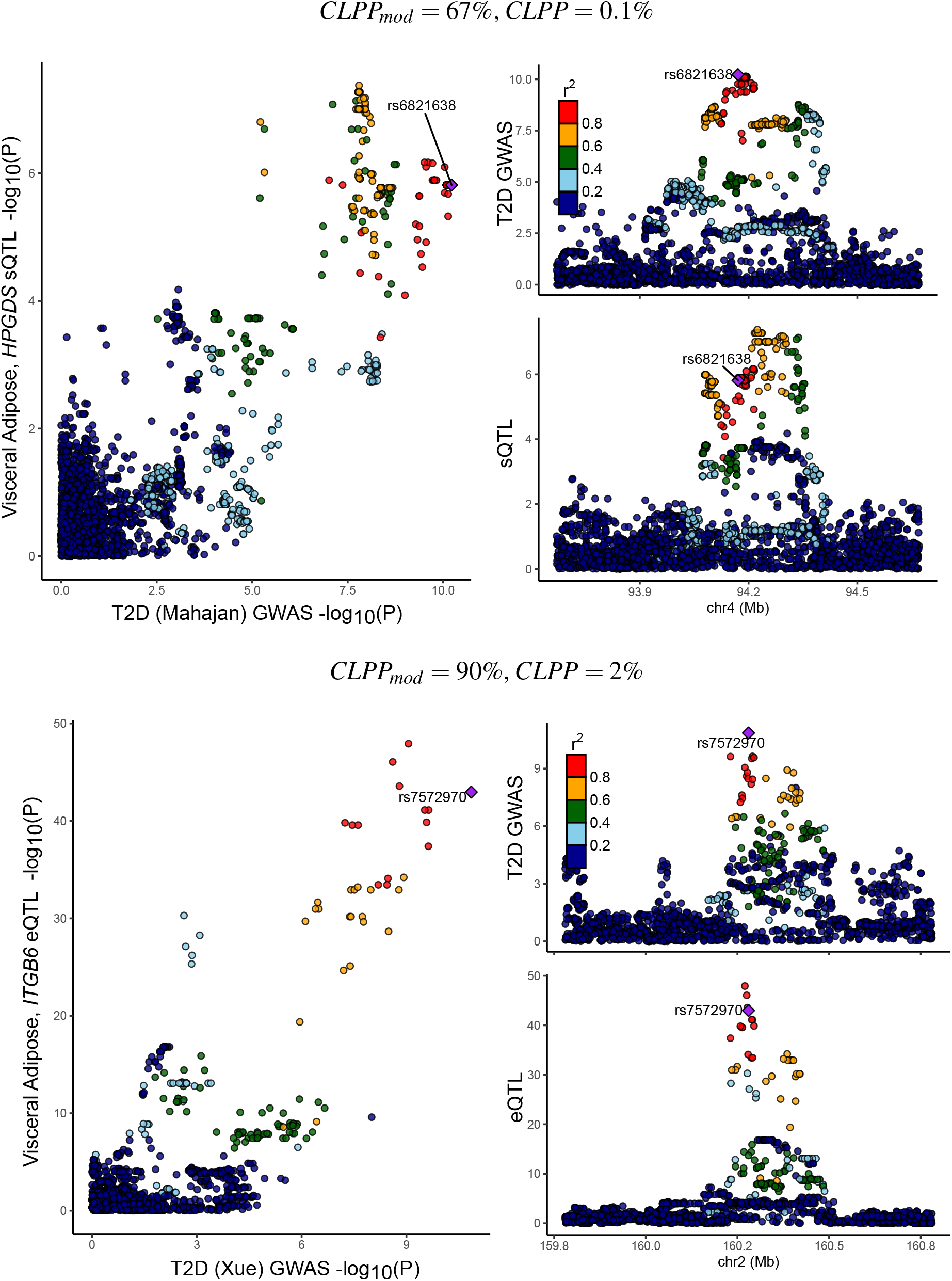
Example LocusCompare plots for each quartile of the CLPP-mod score.

**Figure S4:**
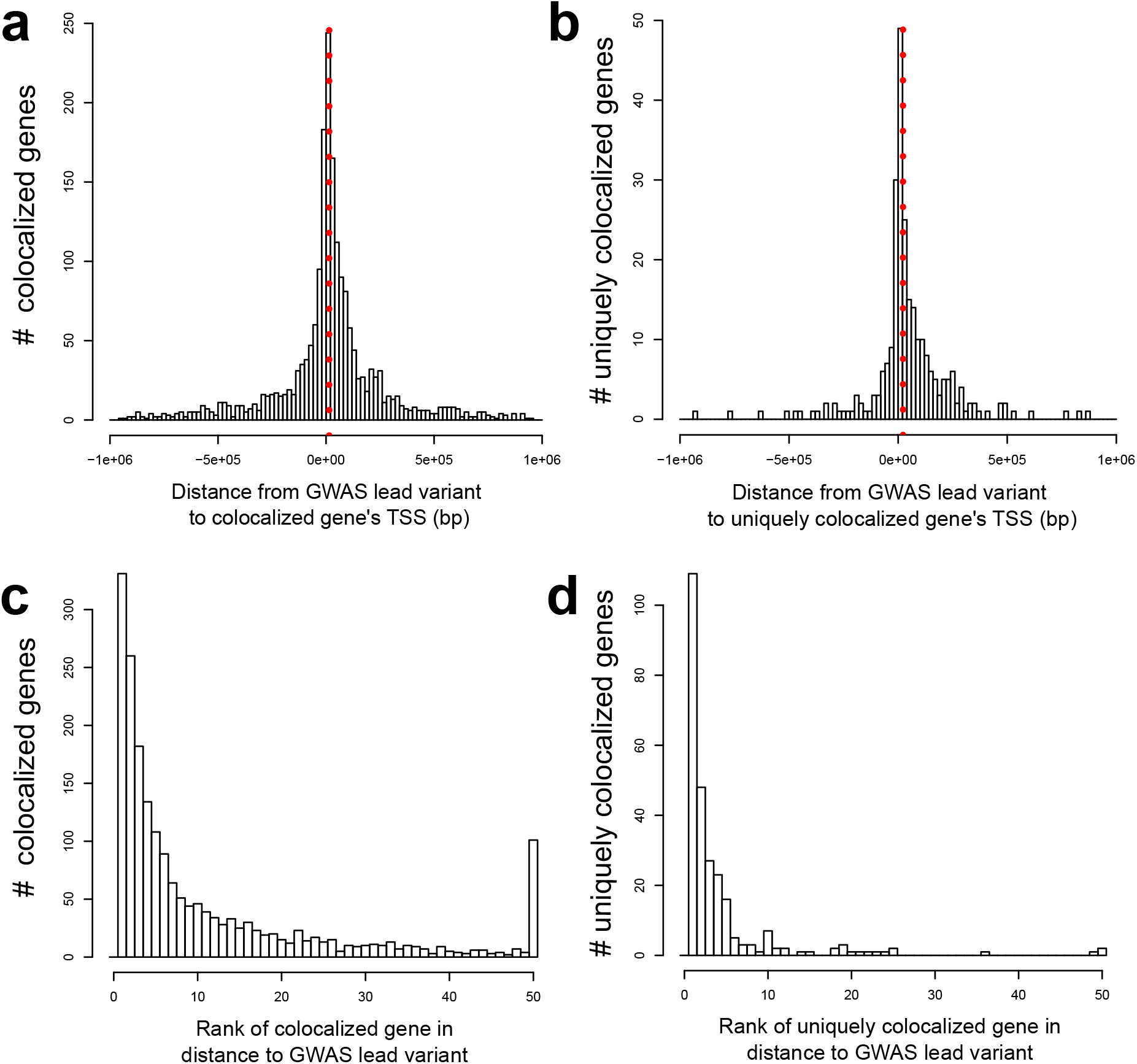
Characteristics of candidate and colocalized genes. (A) Histogram of distance from colocalized genes to the lead GWAS variant. Positive values on the x-axis indicate that a GWAS variant lies upstream of the TSS. (B) Histogram of distance from colocalized genes to the lead GWAS variant at loci with just one colocalizing gene. (C) Proximity (ranked by distance) of colocalized genes’ TSS to lead GWAS variant. A rank of 1 indicates that the colocalizing gene is the closest gene to the GWAS variant. (D) Rank proximity (by distance) of colocalized genes’ TSS to lead GWAS variant at loci with just one colocalizing gene.

**Figure S5:**
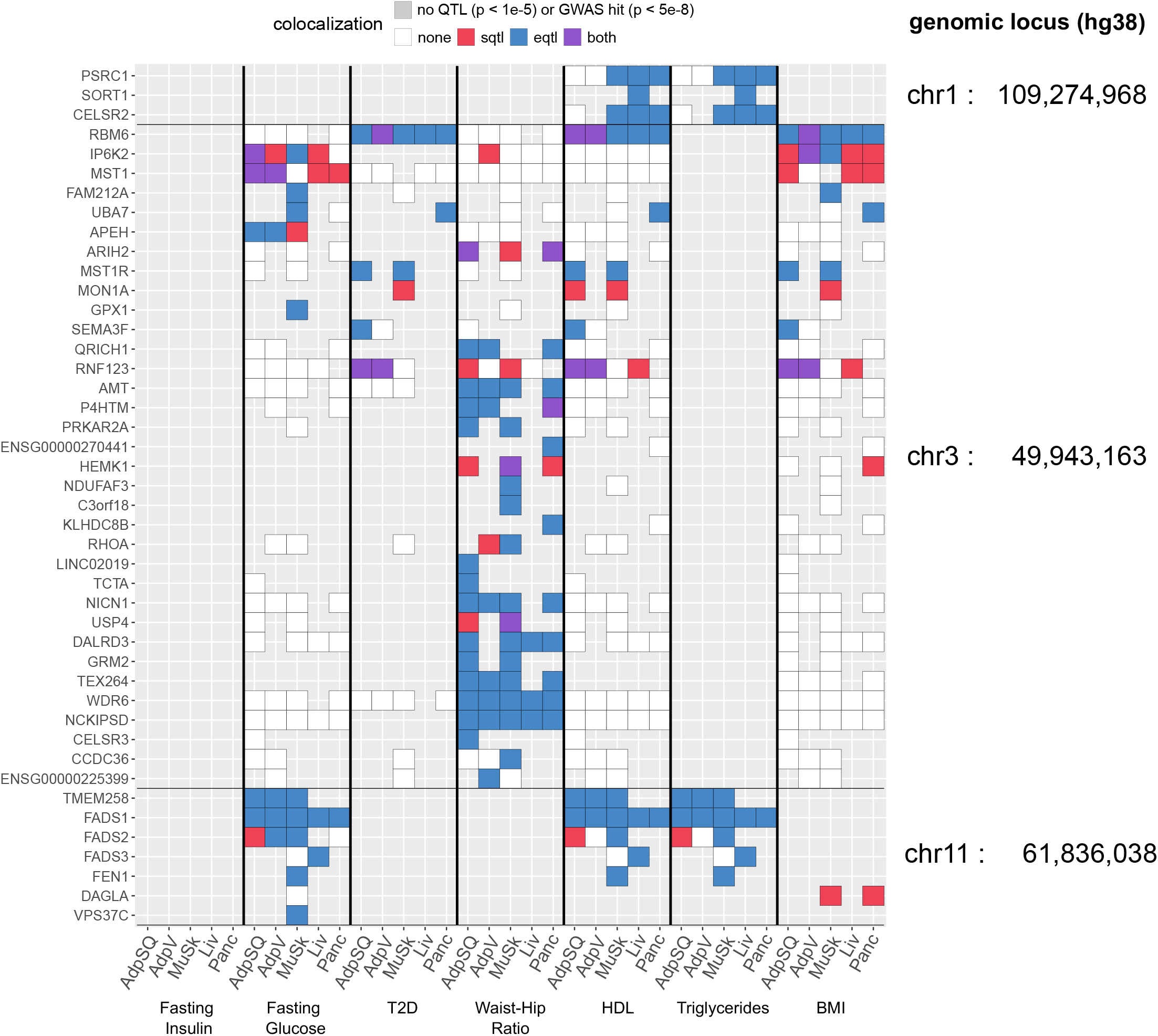
Three separate loci with multiple colocalized genes. Bold horizontal lines separate the genes at each of the three loci.

**Figure S6:**
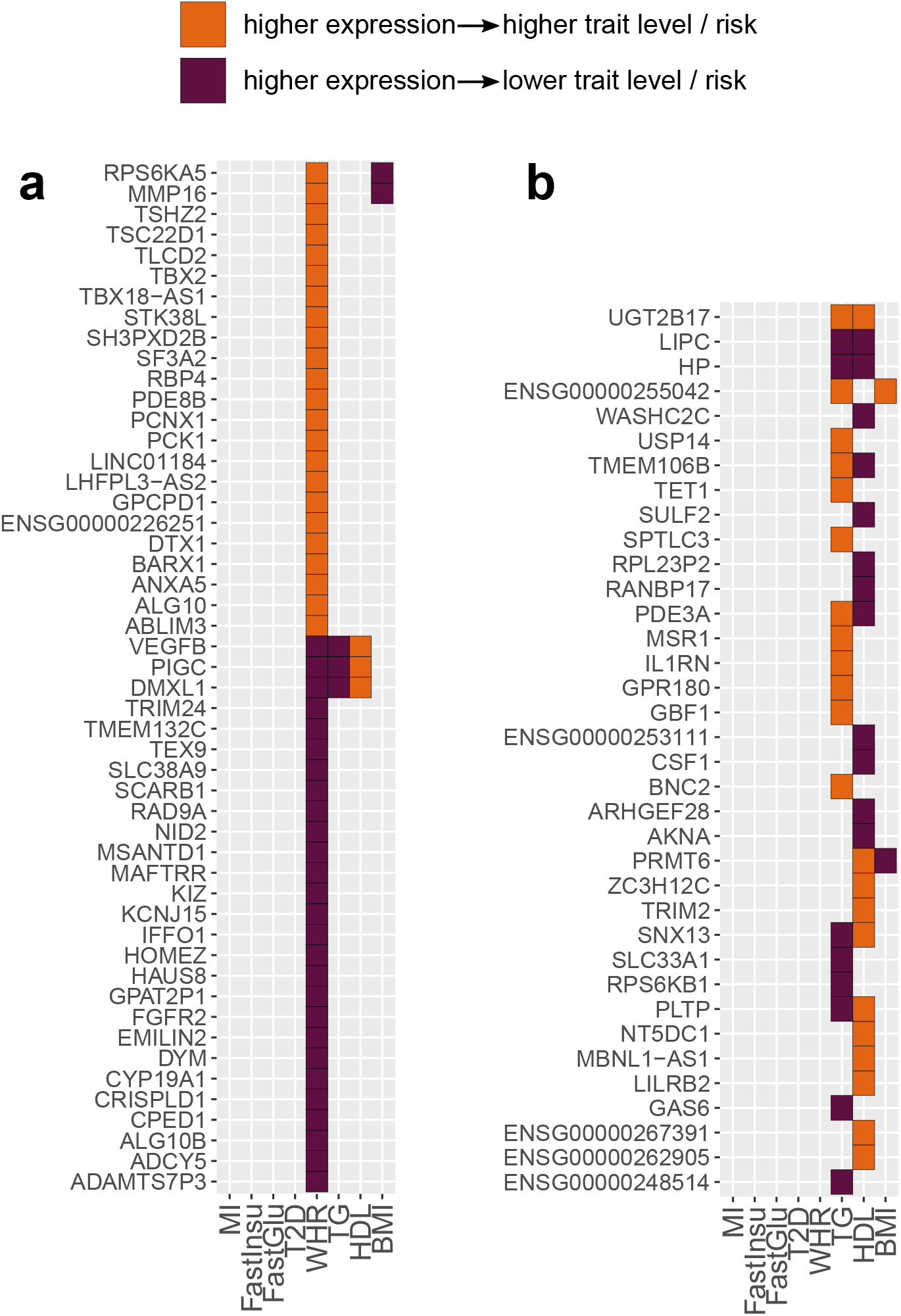
Effects of increased gene expression on cardiometabolic trait level / risk, according to the alignment of GWAS and eQTL directions at colocalized loci. (A) Genes colocalized with eQTLs in WHR, but not in T2D, fasting glucose, fasting insulin, or insulin sensitivity. (B) Genes colocalized with eQTLs in TG and/or HDL but not in WHR, T2D, fasting glucose, fasting insulin, or insulin sensitivity.

**Figure S7:**
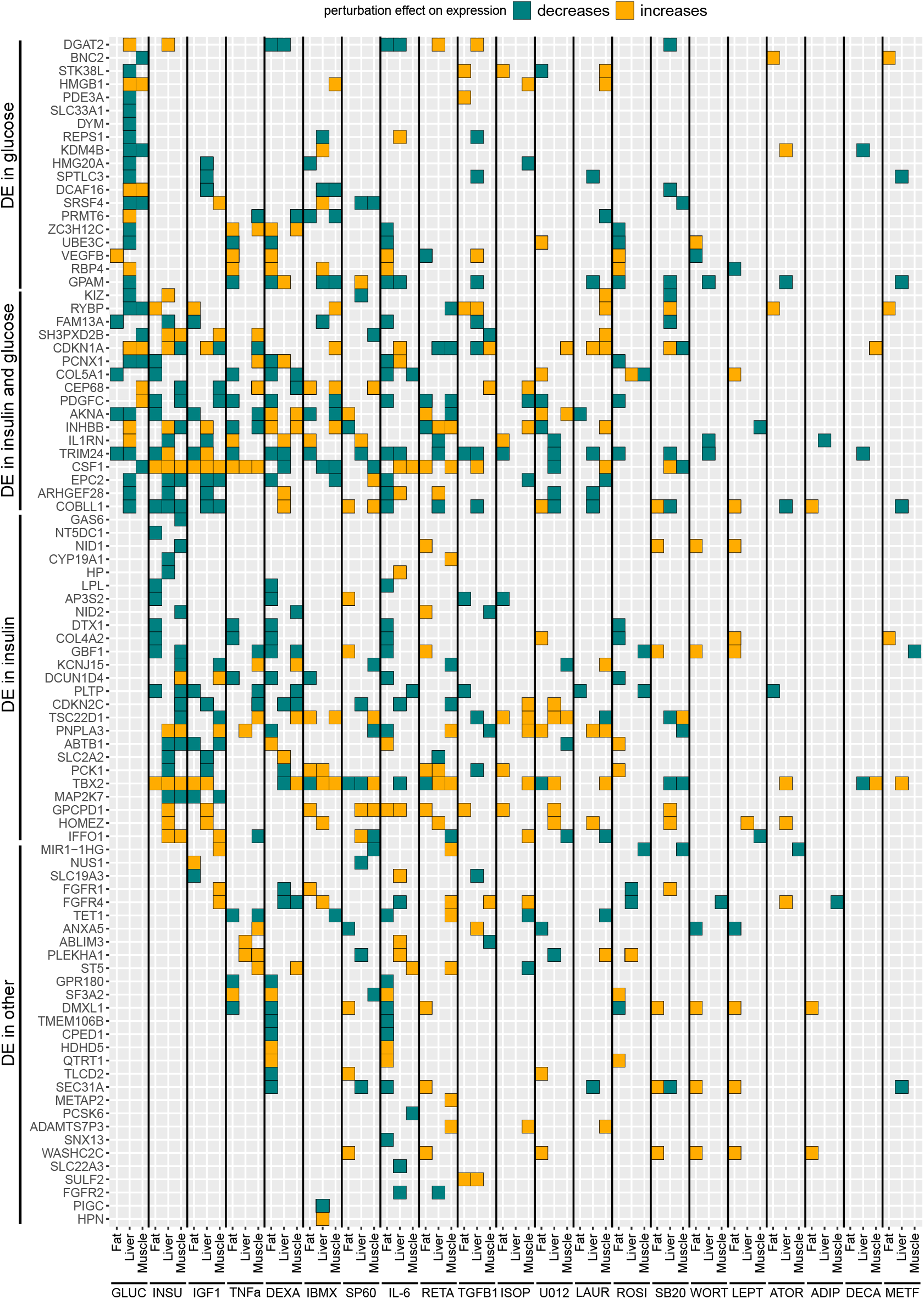
All uniquely colocalized genes that are differentially expressed under at least one perturbation condition.

**Figure S8:**
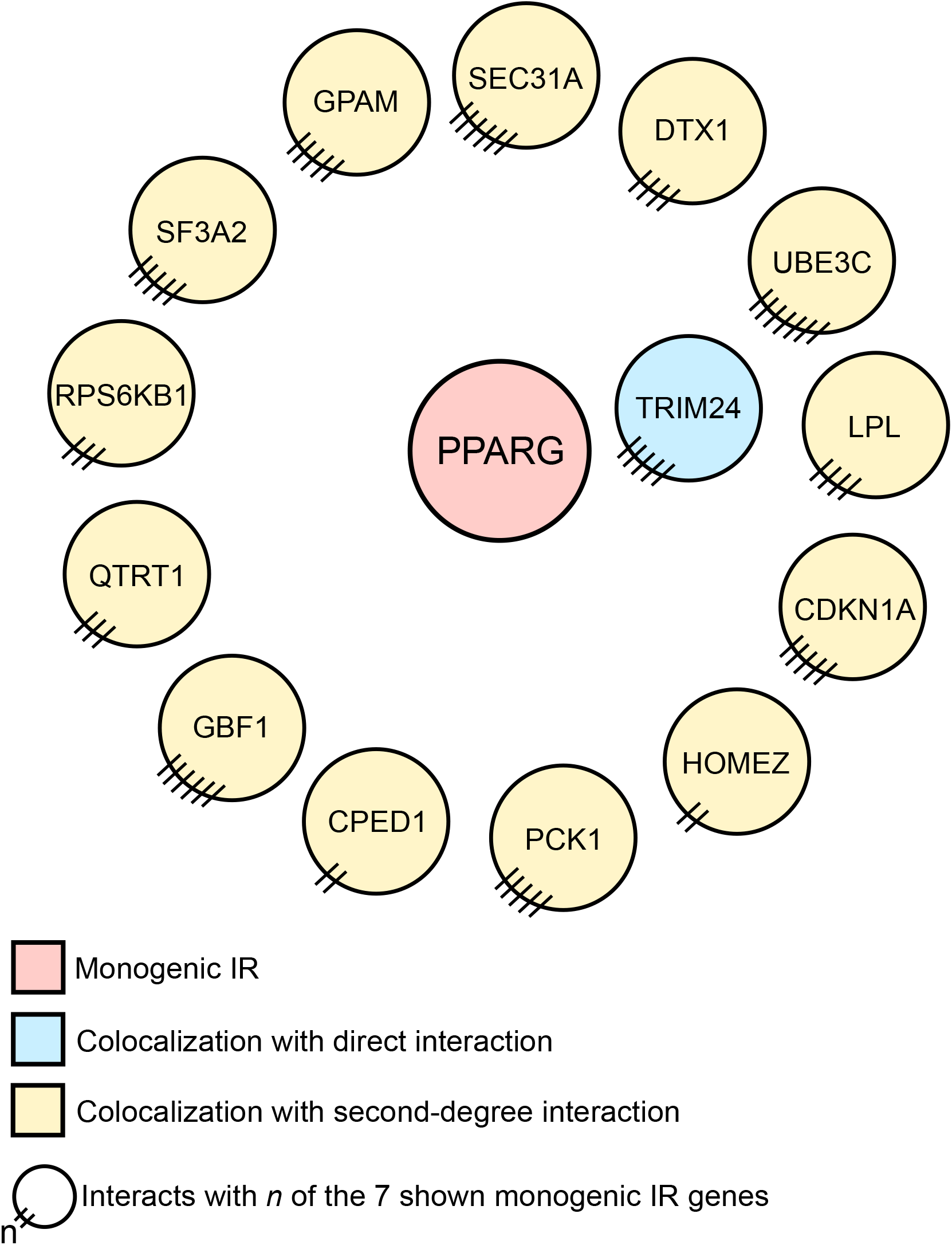
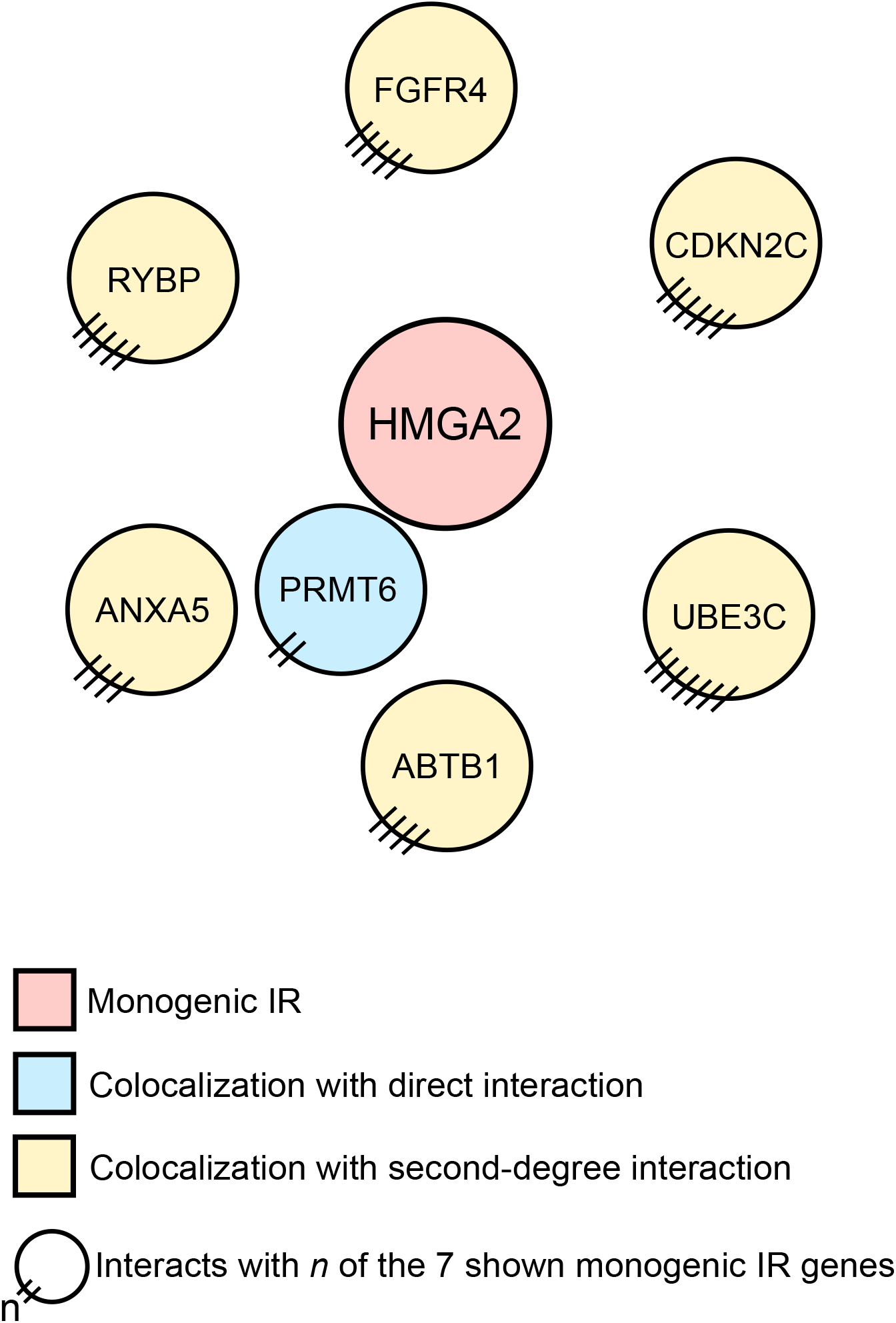
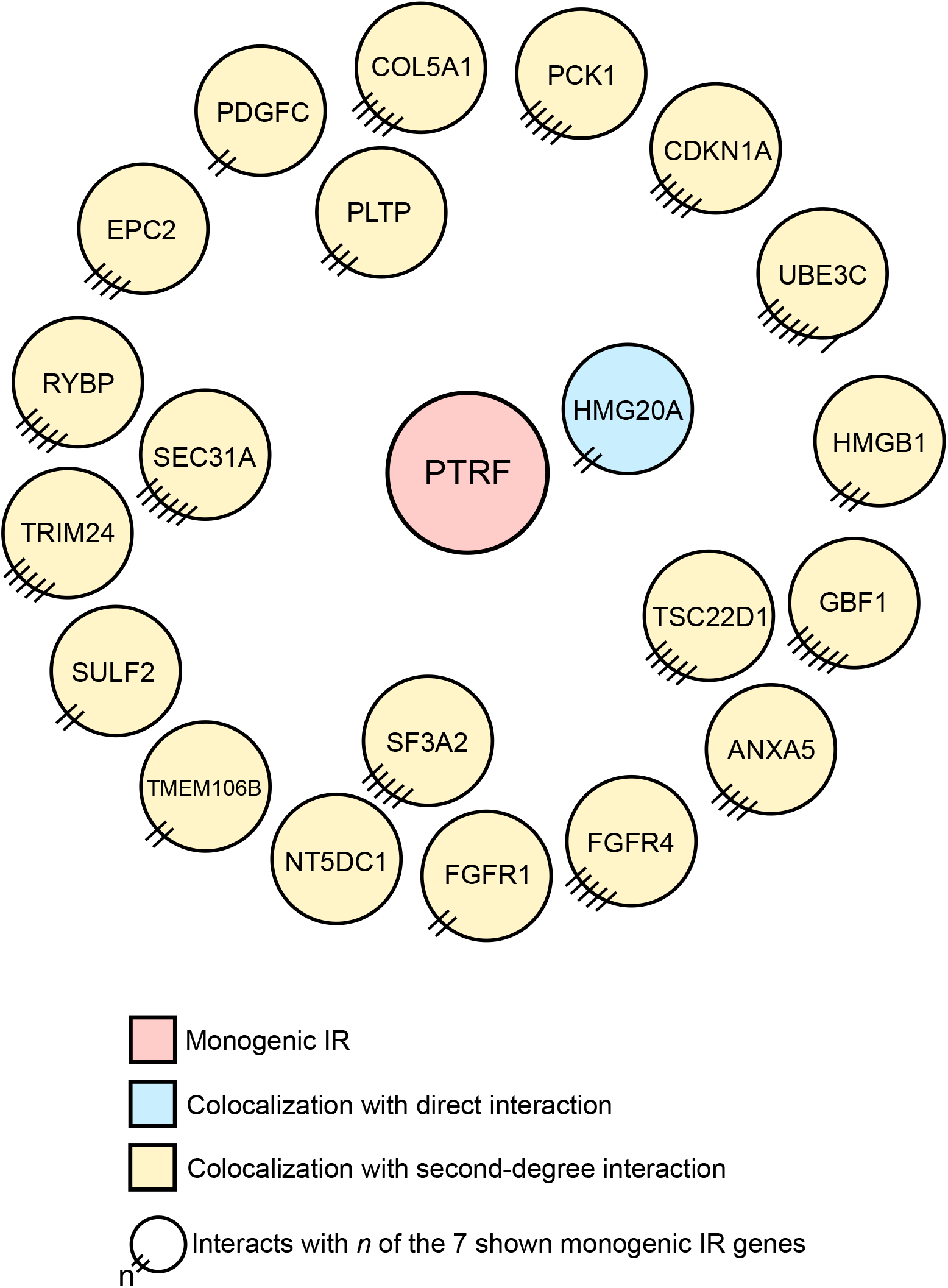
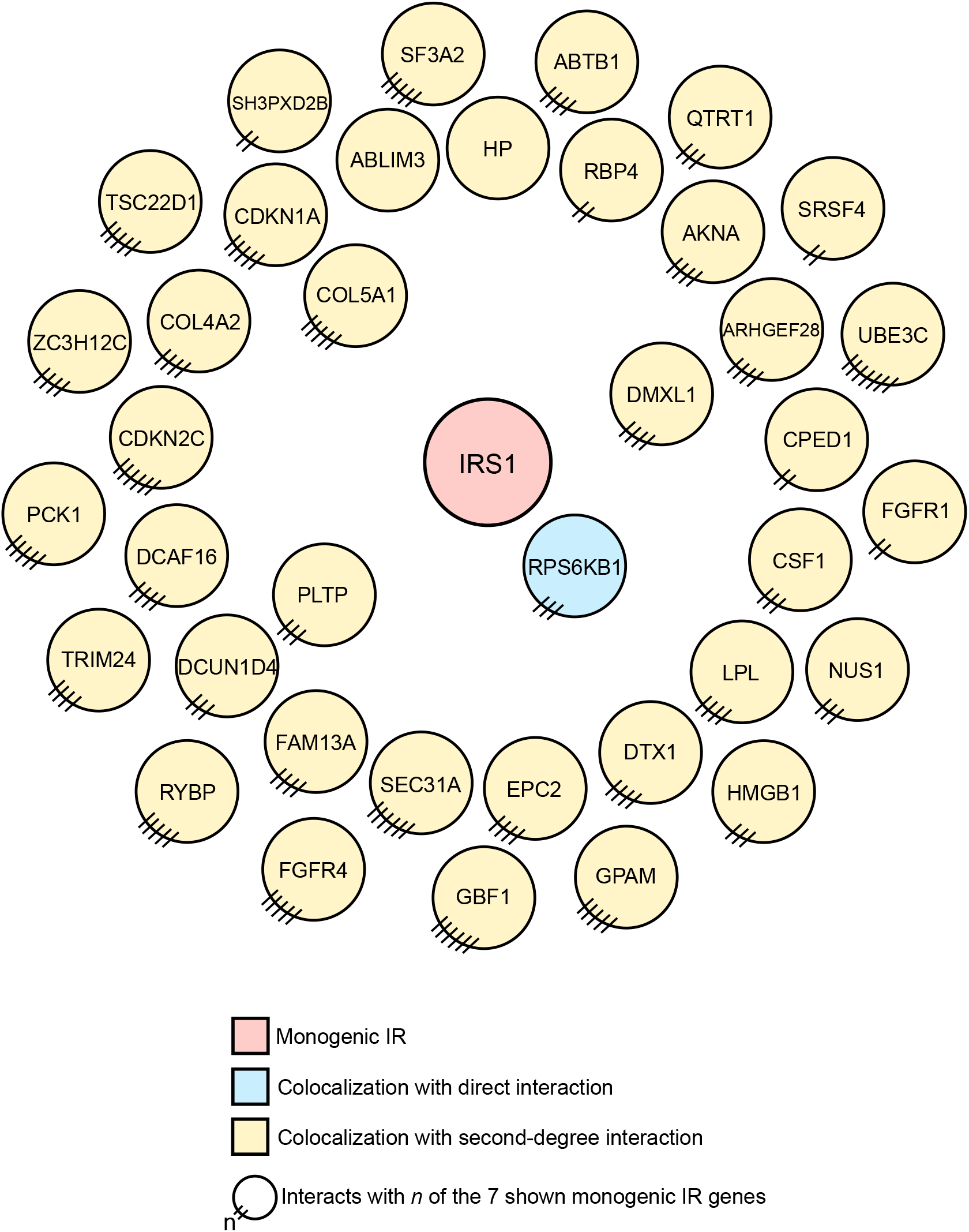
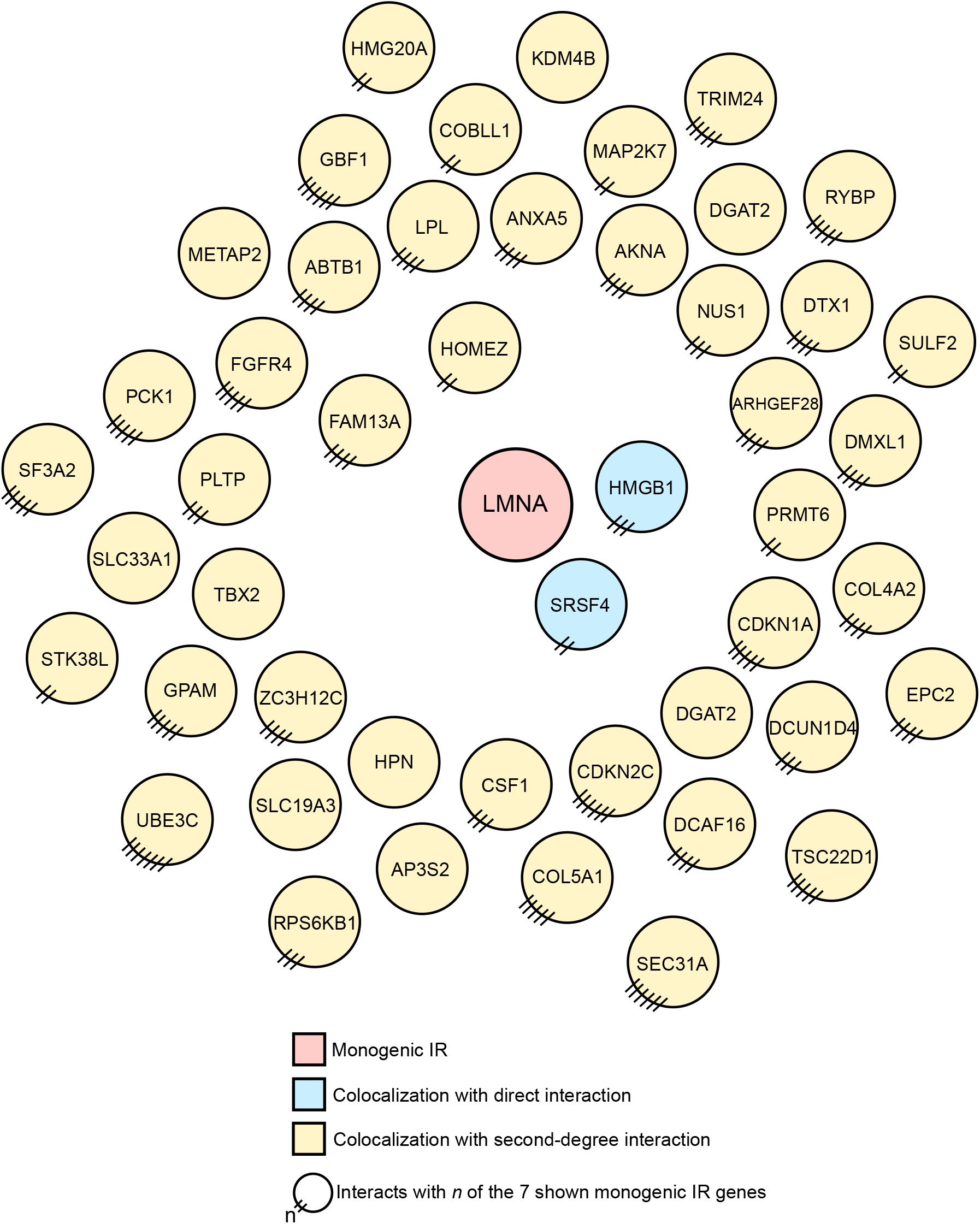
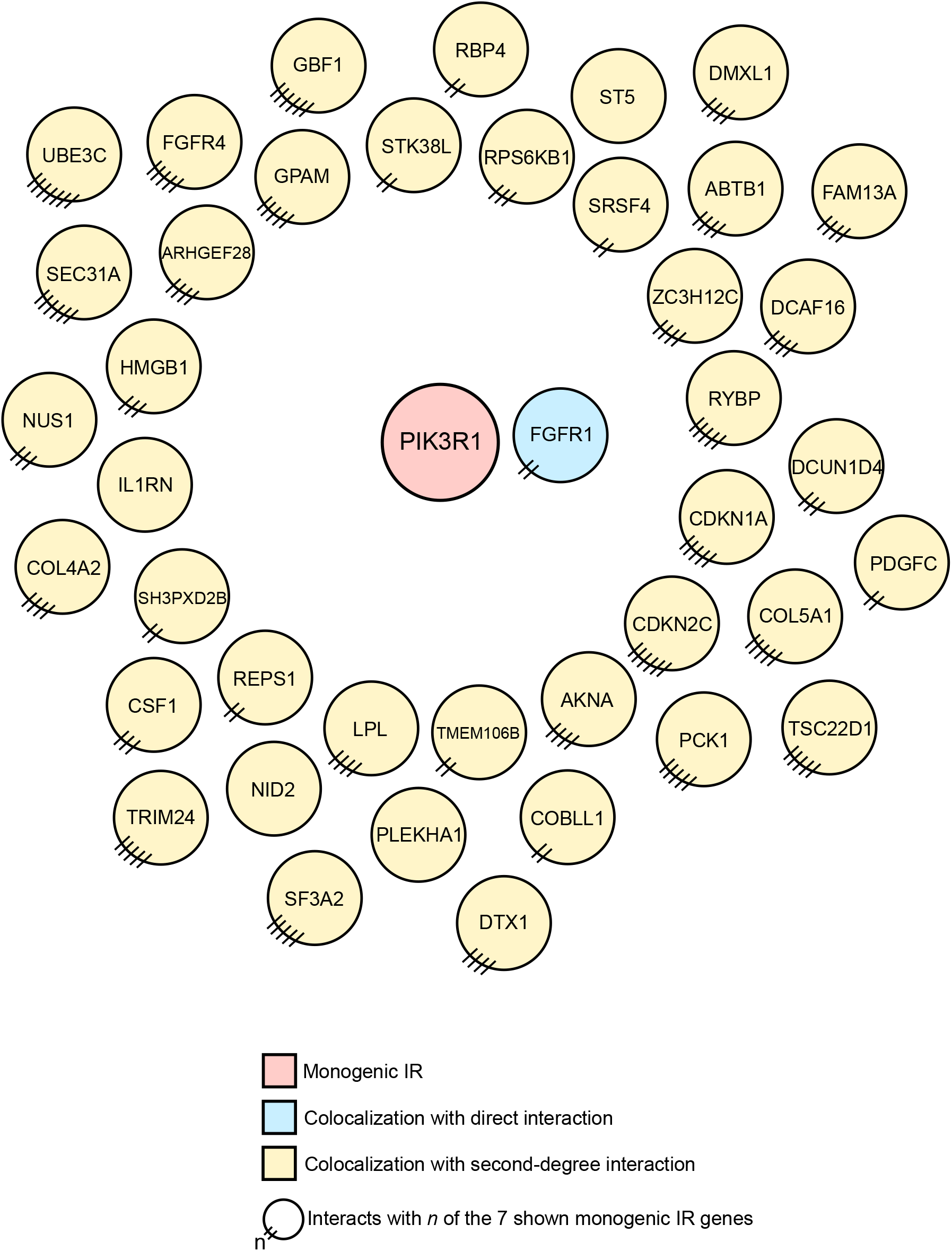
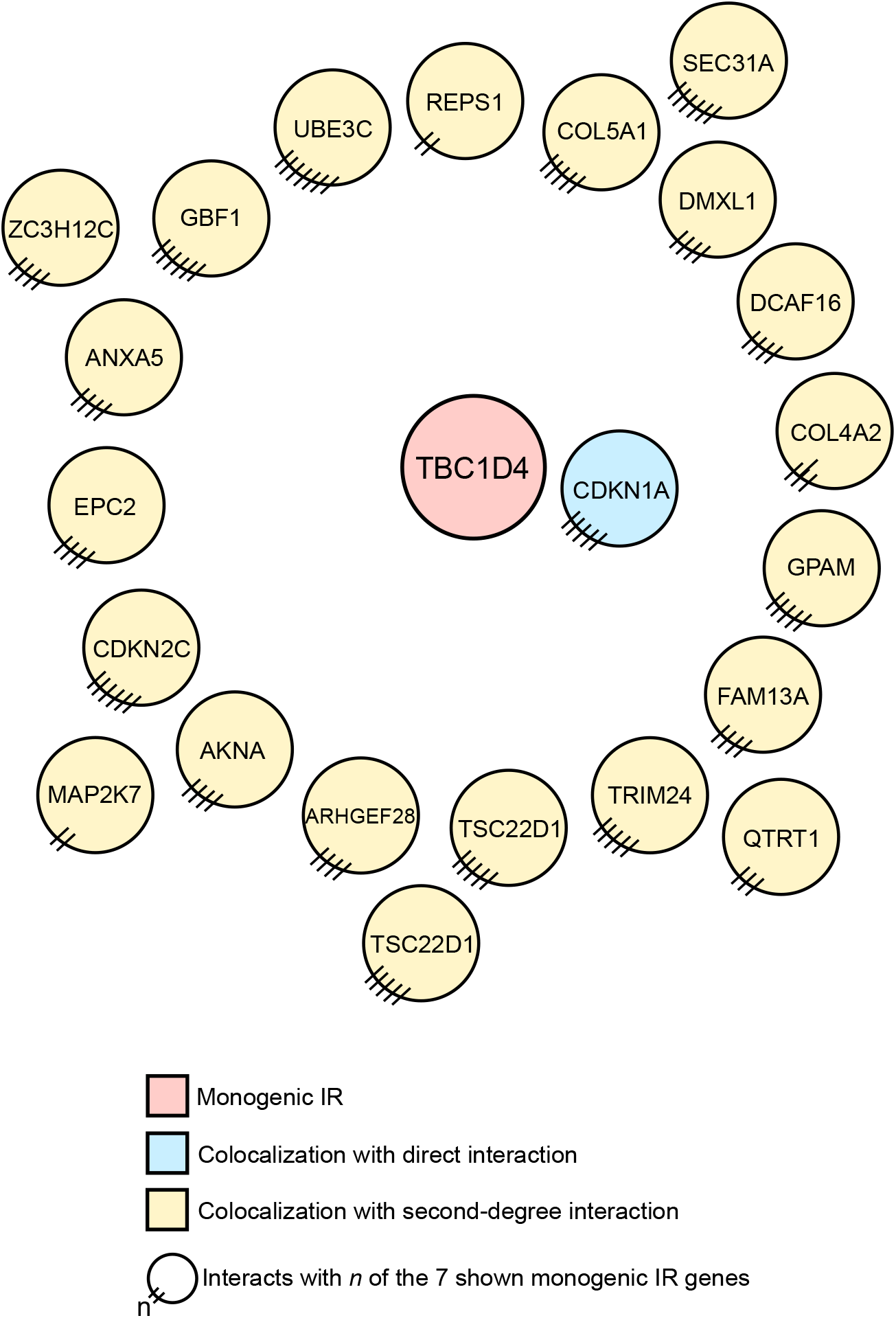
Proximal interaction networks for each of the monogenic IR or T2D genes that directly interacts with at least one of the uniquely colocalized genes in perturbation conditions. All first- and second-degree interactions are shown and color-coded.

**Figure S9:**
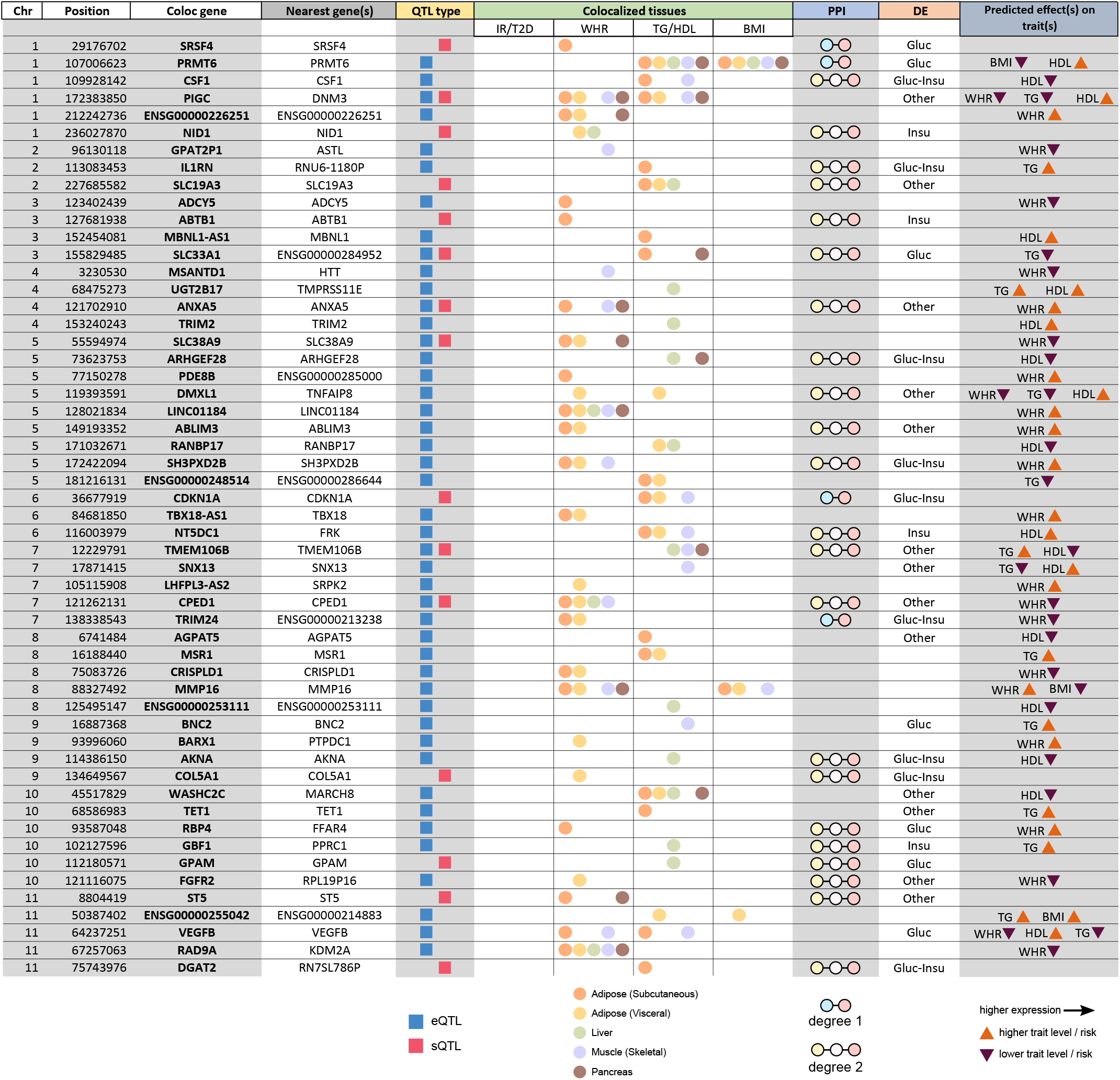
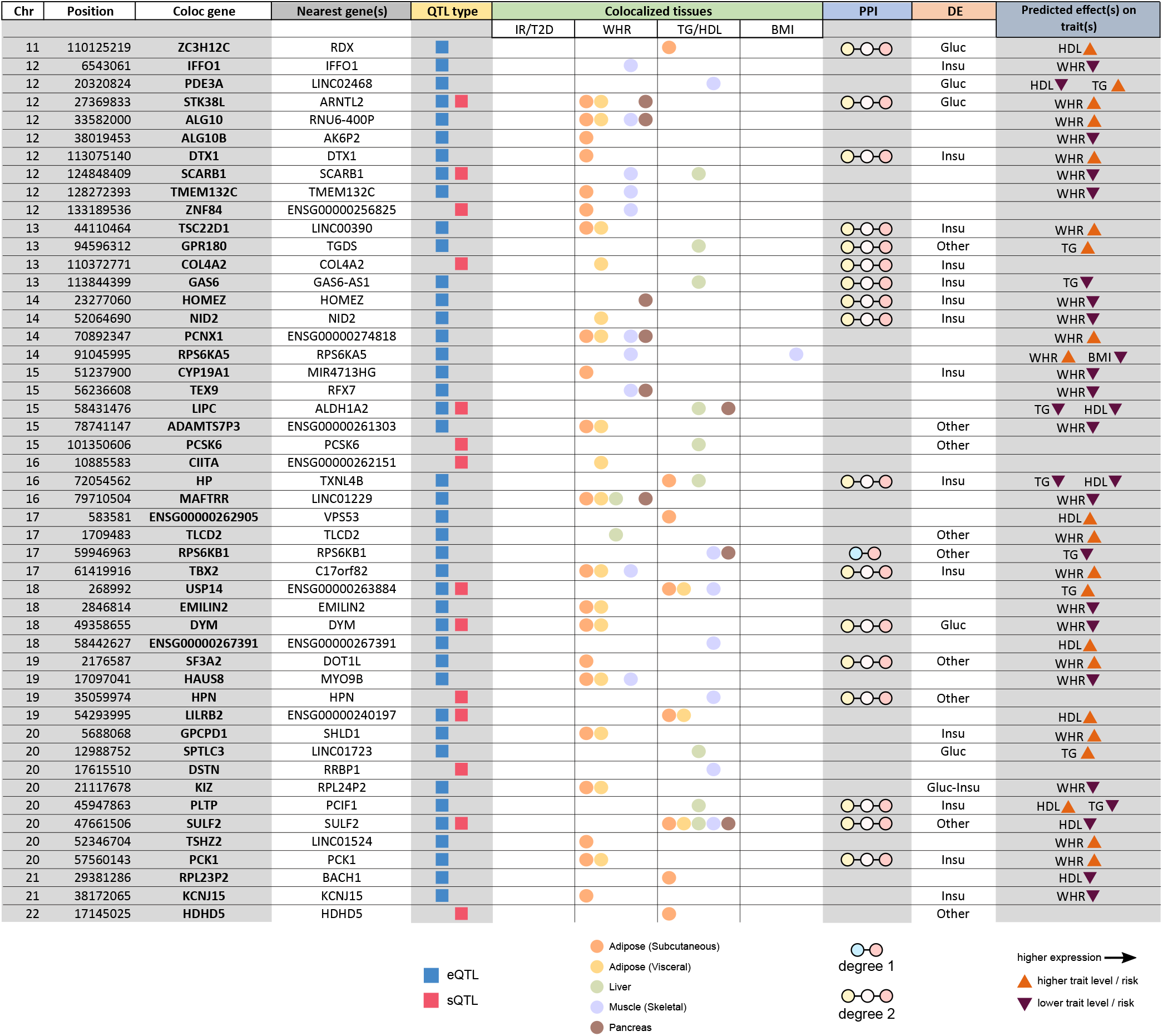
Integrative summary table for all uniquely colocalized genes at loci with a WHR, TG, and/or HDL colocalization, but not an insulin sensitivity, T2D, fasting glucose, or fasting insulin colocalization.

**Table S1:**
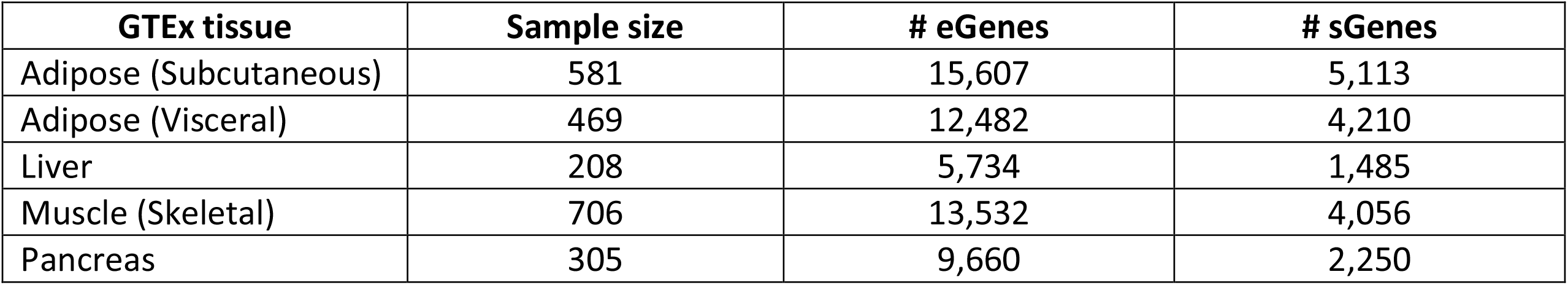
GTEx QTL tissues used for colocalization analysis.

**Table S2:** GWAS used for colocalization analysis.

| Trait | Reference | Ancestry | N_total | N_cases | # hits ($P < 5e-8$) | N of SNPs tested |
| --- | --- | --- | --- | --- | --- | --- |
| Fasting insulin | Scott et al 2012 | European | 108K | N/A | 13 | 64K |
| Fasting glucose | Scott et al 2012 | European | 133K | N/A | 37 | 64K |
| Insulin sensitivity index | Walford et al 2016 | European | 16K | N/A | 10 | 2.4M |
| Insulin sensitivity | Knowles 2015 | Mixed | 2.7K | N/A | 68 | 11M |
| BMI | Yengo et al 2018 | European | 700K | N/A | 1025 | 16M |
| Waist-hip ratio (BMI-adjusted) | Pulit et al 2019 | European | 700K | N/A | 502 | 27M |
| Type 2 diabetes | Mahajan et al 2018 | European | 898K | 74K | 238 | 27M |
| Type 2 diabetes | Suzuki et al 2019 | Japanese | 190K | 36K | 90 | 12M |
| Type 2 diabetes | Spracklen et al 2020 | East Asian | 430K | 77K | 178 | 12M |
| Type 2 diabetes | Xue et al 2018 | European | 650K | 63K | 133 | 16M |
| Triglycerides | Klarin et al 2018 | Mixed | >600K | N/A | 263 | 23M |
| HDL | Klarin et al 2018 | Mixed | 297K | N/A | 302 | 23M |

**Table S3:** Number of candidate/colocalized genes and loci per QTL type / tissue.

| QTL tissue | QTL type | # candidate genes | # candidate loci | # colocalized genes | # colocalized loci |
| --- | --- | --- | --- | --- | --- |
| Adipose (Subcutaneous) | eQTL | 2406 | 611 | 584 | 275 |
| Adipose (Subcutaneous) | sQTL | 1108 | 442 | 281 | 187 |
| Adipose (Subcutaneous) | either | 2972 | 667 | 768 | 330 |
| Adipose (Visceral) | eQTL | 1850 | 552 | 447 | 234 |
| Adipose (Visceral) | sQTL | 974 | 428 | 252 | 168 |
| Adipose (Visceral) | either | 2409 | 628 | 629 | 300 |
| Liver | eQTL | 728 | 334 | 237 | 142 |
| Liver | sQTL | 469 | 278 | 123 | 96 |
| Liver | either | 1089 | 433 | 330 | 190 |
| Muscle (Skeletal) | eQTL | 2154 | 583 | 513 | 241 |
| Muscle (Skeletal) | sQTL | 940 | 428 | 236 | 161 |
| Muscle (Skeletal) | either | 2653 | 642 | 673 | 294 |
| Pancreas | eQTL | 1355 | 467 | 371 | 194 |
| Pancreas | sQTL | 580 | 324 | 148 | 113 |
| Pancreas | either | 1678 | 524 | 458 | 228 |

**Table S4:** Number of candidate/colocalized genes and loci per GWAS. (Note that the numbers of candidate loci are not identical to the numbers in Table S2 because some loci overlap no QTLs and therefore are excluded here.)

| GWAS trait | Study | # candidate genes | # candidate loci | # colocalized genes | # colocalized loci |
| --- | --- | --- | --- | --- | --- |
| Fasting insulin | Scott et al 2012 | 33 | 11 | 17 | 7 |
| Fasting glucose | Scott et al 2012 | 125 | 21 | 36 | 13 |
| Insulin sensitivity index | Walford et al 2016 | 31 | 8 | 9 | 4 |
| Insulin sensitivity | Knowles 2015 | 51 | 16 | 4 | 3 |
| BMI | Yengo et al 2018 | 2421 | 526 | 506 | 223 |
| Waist-hip ratio | Pulit et al 2019 | 1617 | 311 | 395 | 169 |
| Type 2 diabetes | Mahajan et al 2018 | 737 | 166 | 190 | 89 |
| Type 2 diabetes | Suzuki et al 2019 | 201 | 59 | 33 | 22 |
| Type 2 diabetes | Spracklen et al 2020 | 427 | 113 | 92 | 49 |
| Type 2 diabetes | Xue et al 2018 | 378 | 95 | 118 | 52 |
| Triglycerides | Klarin et al 2018 | 1105 | 187 | 287 | 111 |
| HDL | Klarin et al 2018 | 1204 | 210 | 263 | 123 |

**Table S5:** Fraction of tissue-specific, single-gene colocalizations in each tissue / disease combination. Each cell represents the percentage of a trait category’s tissue-specific genes that were found in a given tissue. Heavier shading indicates a higher contribution of the tissue to that disease category than to other disease categories. Some tissue-specific loci contribute to more than one disease. Each column sums to 100% (with small deviation due to rounding error).

|  | T2D, IS | WHR | TG, HDL |
| --- | --- | --- | --- |
| Adipose | 50% | 79% | 44% |
| Muscle | 24% | 14% | 21% |
| Liver | 15% | 5% | 36% |
| Pancreas | 12% | 2% | 0% |

**Table S6:**
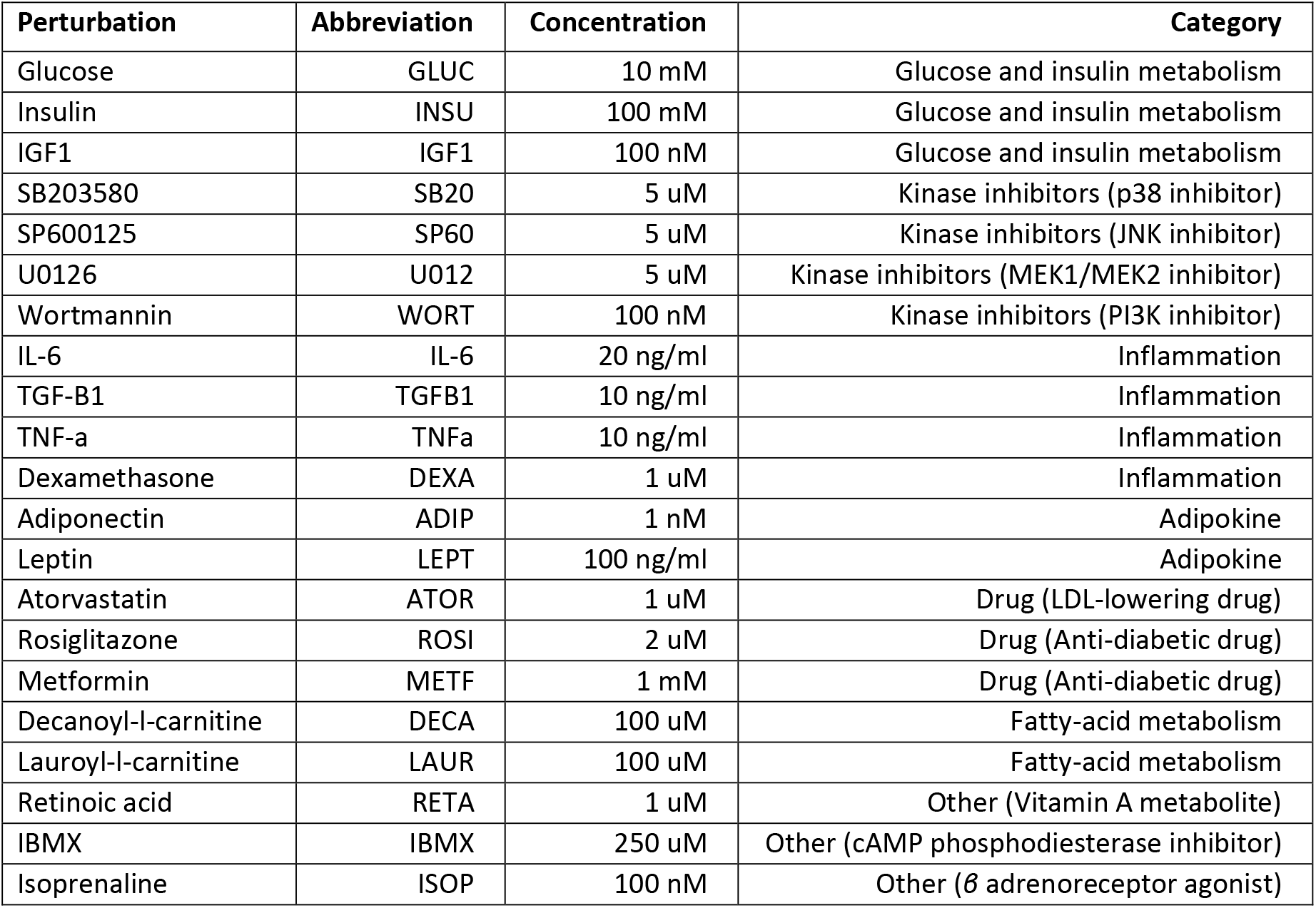
List of metabolic perturbations and their abbreviations.

**Table S7:** Genetic studies-based IR/T2D genes used in PPI network analysis.

| Gene | Source |
| --- | --- |
| *ANKRD55* | Bonnefond [60] |
| *ARL15* | Bonnefond, Brown [61] |
| *FTO* | Bonnefond, Brown, Prasad [62] |
| *GCKR* | Bonnefond, Brown |
| *GRB14* | Bonnefond |
| *HMGA2* | Bonnefond |
| *IGF1* | Brown, Prasad |
| *IRS1* | Brown, Prasad |
| *KLF14* | Bonnefond, Brown, Prasad |
| *LEP* | Bonnefond |
| *MC4R* | Bonnefond, Brown, Prasad |
| *MSMO1* | Prasad |
| *NAT2* | Brown |
| *PEPD* | Bonnefond |
| *PPARG* | Bonnefond, Brown |
| *RBMS1* | Bonnefond |
| *SC4MOL* | Brown |
| *SLC16A11* | Bonnefond |
| *TCERG1L* | Brown, Prasad |
| *TCF7L2* | Brown |
| *TMEM163* | Brown, Prasad |

**Table S8:**
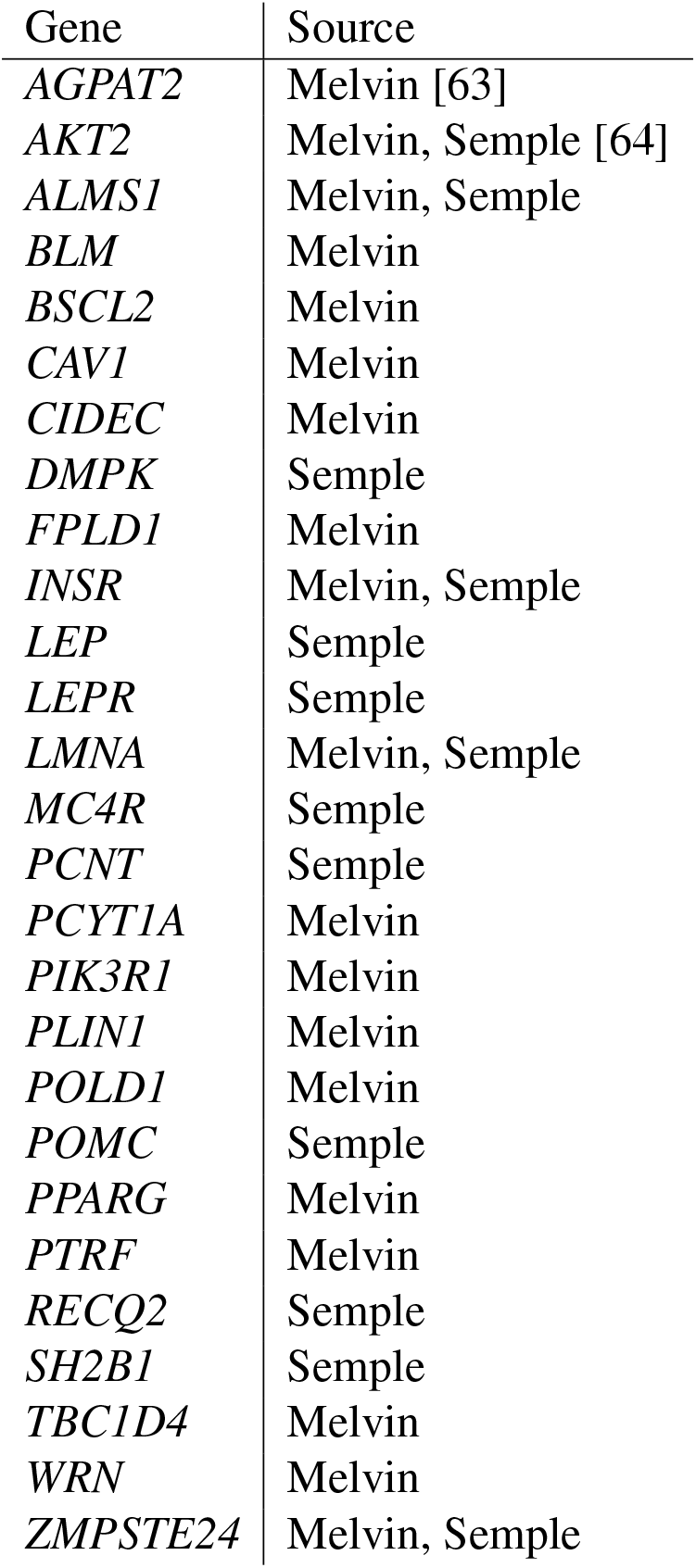
Monogenic IR/T2D genes used in PPI network analysis.

## Additional Data Files

**Additional Data File S1: All GWAS-SNP associations, annotated with the corresponding locus numbers.**

**Additional Data File S2: Index of all genomic loci, with chromosomal coordinates (hg38), determined using LDetect.**

**Additional Data File S3: All colocalization test results.** A colocalization test applies to a unique combination of a GWAS trait, a QTL type (eQTL or sQTL), a QTL feature (gene expression level or intragenic splice junction), a QTL tissue, and a lead GWAS SNP.

**Additional Data File S4: Categorization of loci based on number of candidate and colocalized genes, tissue specificity, and most relevant colocalized traits.**

**Additional Data File S5: Cell type-specificity for uniquely colocalized genes according to single-cell analysis.** Note that a gene can be specific to more than just one cell type.

**Additional Data File S6: GTEx-inferred WGCNA modules associated with uniquely colocalized genes, along with gene set enrichment tests for these modules.**

**Additional Data File S7: Directional effects of gene expression on GWAS traits for all colocalized gene / eQTL tissue / GWAS trait combinations.**

**Additional Data File S8: Effect directions of metabolic perturbations on expression levels of colocalized genes.**

**Additional Data File S9: Protein-protein interactions between known monogenic diabetes/IR genes and uniquely colocalized genes, determined by co-regulation under metabolic perturbations.**

**Additional Data File S10: Protein-protein interactions of uniquely colocalized protein-coding genes under 21 metabolic perturbations each in fat, muscle, and liver cells.**

## References

[1] Mahajan A, Taliun D, Thurner M, Robertson NR, Torres JM, Rayner NW, et al. Fine-mapping type 2 diabetes loci to single-variant resolution using high-density imputation and islet-specific epigenome maps. Nature Genetics 2018;50(11):1505–1513. doi:10.1038/s41588-018-0241-6.

[2] Spracklen CN, Horikoshi M, Kim YJ, Lin K, Bragg F, Moon S, et al. Identification of type 2 diabetes loci in 433,540 East Asian individuals. Nature 2020;582(7811):240–245. doi:10.1038/s41586-020-2263-3.

[3] Suzuki K, Akiyama M, Ishigaki K, Kanai M, Hosoe J, Shojima N, et al. Identification of 28 new susceptibility loci for type 2 diabetes in the Japanese population. Nature Genetics 2019; 51(3):379–386. doi:10.1038/s41588-018-0332-4.

[4] Xue A, Wu Y, Zhu Z, Zhang F, Kemper KE, Zheng Z, et al. Genome-wide association analyses identify 143 risk variants and putative regulatory mechanisms for type 2 diabetes. Nature Communications 2018;9(1):2941. doi:10.1038/s41467-018-04951-w.

[5] Mahajan A, Wessel J, Willems SM, Zhao W, Robertson NR, Chu AY, et al. Refining the accuracy of validated target identification through coding variant fine-mapping in type 2 diabetes. Nature Genetics 2018;50(4):559–571. doi:10.1038/s41588-018-0084-1. Number: 4 Publisher: Nature Publishing Group.

[6] Udler MS, Kim J, von Grotthuss M, Bonàs-Guarch S, Cole JB, Chiou J, et al. Type 2 diabetes genetic loci informed by multi-trait associations point to disease mechanisms and subtypes: A soft clustering analysis. PLoS Medicine 2018;15(9). doi:10.1371/journal.pmed.1002654.

[7] Wagner R, Heni M, Tabàk AG, Machann J, Schick F, Randrianarisoa E, et al. Pathophysiology-based subphenotyping of individuals at elevated risk for type 2 diabetes. Nature Medicine 2021;27(1):49–57. doi:10.1038/s41591-020-1116-9. Number: 1 Publisher: Nature Publishing Group.

[8] Zaharia OP, Strassburger K, Strom A, Bönhof GJ, Karusheva Y, Antoniou S, et al. Risk of diabetes-associated diseases in subgroups of patients with recent-onset diabetes: a 5-year follow-up study. The Lancet Diabetes & Endocrinology 2019;7(9):684–694. doi:10.1016/S2213-8587(19)30187-1.

[9] Ahlqvist E, Storm P, Kääjämäki A, Martinell M, Dorkhan M, Carlsson A, et al. Novel subgroups of adult-onset diabetes and their association with outcomes: a data-driven cluster analysis of six variables. The Lancet Diabetes & Endocrinology 2018;6(5):361–369. doi:10.1016/S2213-8587(18)30051-2.

[10] Lauro D, Kido Y, Castle AL, Zarnowski MJ, Hayashi H, Ebina Y, Accili D. Impaired glucose tolerance in mice with a targeted impairment of insulin action in muscle and adipose tissue. Nature Genetics 1998;20(3):294–298. doi:10.1038/3112. Number: 3 Publisher: Nature Publishing Group.

[11] Perry JRB, Frayling TM. New gene variants alter type 2 diabetes risk predominantly through reduced beta-cell function. Current Opinion in Clinical Nutrition and Metabolic Care 2008; 11(4):371–377. doi:10.1097/MCO.0b013e32830349a1.

[12] Florez JC. Newly identified loci highlight beta cell dysfunction as a key cause of type 2 diabetes: where are the insulin resistance genes? Diabetologia 2008;51(7):1100–1110. doi:10.1007/s00125-008-1025-9.

[13] Strawbridge RJ, Dupuis J, Prokopenko I, Barker A, Ahlqvist E, Rybin D, et al. Genome-wide association identifies nine common variants associated with fasting proinsulin levels and provides new insights into the pathophysiology of type 2 diabetes. Diabetes 2011;60(10):2624– 2634. doi:10.2337/db11-0415.

[14] Singh R, De Aguiar RB, Naik S, Mani S, Ostadsharif K, Wencker D, Sotoudeh M, Malekzadeh R, Sherwin RS, Mani A. LRP6 enhances glucose metabolism by promoting TCF7L2-dependent insulin receptor expression and IGF receptor stabilization in humans. Cell Metabolism 2013;17(2):197–209. doi:10.1016/j.cmet.2013.01.009.

[15] Zhang J, McKenna LB, Bogue CW, Kaestner KH. The diabetes gene Hhex maintains *δ*-cell differentiation and islet function. Genes & Development 2014;28(8):829–834. doi:10.1101/gad.235499.113.

[16] Gao T, McKenna B, Li C, Reichert M, Nguyen J, Singh T, et al. Pdx1 maintains *β* cell identity and function by repressing an *α* cell program. Cell Metabolism 2014;19(2):259– 271. doi:10.1016/j.cmet.2013.12.002.

[17] Knowles JW, Xie W, Zhang Z, Chennamsetty I, Chennemsetty I, Assimes TL, et al. Identification and validation of N-acetyltransferase 2 as an insulin sensitivity gene. The Journal of Clinical Investigation 2015;125(4):1739–1751. doi:10.1172/JCI74692.

[18] Walford GA, Gustafsson S, Rybin D, Stančáková A, Chen H, Liu CT, et al. Genome-Wide Association Study of the Modified Stumvoll Insulin Sensitivity Index Identifies BCL2 and FAM19A2 as Novel Insulin Sensitivity Loci. Diabetes 2016;65(10):3200–3211. doi:10.2337/db16-0199.

[19] Rung J, Cauchi S, Albrechtsen A, Shen L, Rocheleau G, Cavalcanti-Proença C, et al. Genetic variant near IRS1 is associated with type 2 diabetes, insulin resistance and hyperinsulinemia. Nature Genetics 2009;41(10):1110–1115. doi:10.1038/ng.443.

[20] Keramati AR, Fathzadeh M, Go GW, Singh R, Choi M, Faramarzi S, et al. A form of the metabolic syndrome associated with mutations in DYRK1B. The New England Journal of Medicine 2014;370(20):1909–1919. doi:10.1056/NEJMoa1301824.

[21] Moltke I, Grarup N, Jørgensen ME, Bjerregaard P, Treebak JT, Fumagalli M, et al. A common Greenlandic TBC1D4 variant confers muscle insulin resistance and type 2 diabetes. Nature 2014;512(7513):190–193. doi:10.1038/nature13425. Number: 7513 Publisher: Nature Publishing Group.

[22] Albert JS, Yerges-Armstrong LM, Horenstein RB, Pollin TI, Sreenivasan UT, Chai S, et al. Null Mutation in Hormone-Sensitive Lipase Gene and Risk of Type 2 Diabetes. New England Journal of Medicine 2014;370(24):2307–2315. doi:10.1056/NEJMoa1315496. Publisher: Massachusetts Medical Society eprint: https://doi.org/10.1056/NEJMoa1315496.

[23] Batista TM, Haider N, Kahn CR. Defining the underlying defect in insulin action in type 2 diabetes. Diabetologia 2021;64(5):994–1006. doi:10.1007/s00125-021-05415-5.

[24] Berisa T, Pickrell JK. Approximately independent linkage disequilibrium blocks in human populations. Bioinformatics (Oxford, England) 2016;32(2):283–285. doi:10.1093/bioinformatics/btv546.

[25] Zhu Z, Zhang F, Hu H, Bakshi A, Robinson MR, Powell JE, et al. Integration of summary data from GWAS and eQTL studies predicts complex trait gene targets. Nature Genetics 2016;48(5):481–487. doi:10.1038/ng.3538. Number: 5 Publisher: Nature Publishing Group.

[26] Glaser C, Heinrich J, Koletzko B. Role of FADS1 and FADS2 polymorphisms in polyunsaturated fatty acid metabolism. Metabolism 2010;59(7):993–999. doi:10.1016/j.metabol.2009.10.022.

[27] Musunuru K, Strong A, Frank-Kamenetsky M, Lee NE, Ahfeldt T, Sachs KV, et al. From noncoding variant to phenotype via SORT1 at the 1p13 cholesterol locus. Nature 2010; 466(7307):714–719. doi:10.1038/nature09266. Number: 7307 Publisher: Nature Publishing Group.

[28] Torres JM, Abdalla M, Payne A, Fernandez-Tajes J, Thurner M, Nylander V, Gloyn AL, Mahajan A, McCarthy MI. A Multi-omic Integrative Scheme Characterizes Tissues of Action at Loci Associated with Type 2 Diabetes. The American Journal of Human Genetics 2020; 107(6):1011–1028. doi:10.1016/j.ajhg.2020.10.009.

[29] Wullschleger S, Wasserman DH, Gray A, Sakamoto K, Alessi DR. Role of TAPP1 and TAPP2 adaptor binding to PtdIns(3,4)P2 in regulating insulin sensitivity defined by knock-in analysis. The Biochemical Journal 2011;434(2):265–274. doi:10.1042/BJ20102012.

[30] Balliu B, Carcamo Orive I, Gloudemans MJ, Nachun DC, Durrant MG, Gazal S, et al. An integrated approach to identify environmental modulators of genetic risk factors for complex traits. bioRxiv 2021;page 2021.02.23.432608. doi:10.1101/2021.02.23.432608. Publisher: Cold Spring Harbor Laboratory Section: New Results.

[31] Han X, Zhou Z, Fei L, Sun H, Wang R, Chen Y, et al. Construction of a human cell landscape at single-cell level. Nature 2020;581(7808):303–309. doi:10.1038/s41586-020-2157-4.

[32] Dougherty JD, Schmidt EF, Nakajima M, Heintz N. Analytical approaches to RNA profiling data for the identification of genes enriched in specific cells. Nucleic Acids Research 2010; 38(13):4218–4230. doi:10.1093/nar/gkq130.

[33] Xu X, Wells AB, O’Brien DR, Nehorai A, Dougherty JD. Cell type-specific expression analysis to identify putative cellular mechanisms for neurogenetic disorders. The Journal of Neuroscience: The Official Journal of the Society for Neuroscience 2014;34(4):1420–1431. doi:10.1523/JNEUROSCI.4488-13.2014.

[34] Langfelder P, Horvath S. WGCNA: an R package for weighted correlation network analysis. BMC Bioinformatics 2008;9(1):559. doi:10.1186/1471-2105-9-559.

[35] de Goede OM, Nachun DC, Ferraro NM, Gloudemans MJ, Rao AS, Smail C, et al. Population-scale tissue transcriptomics maps long non-coding RNAs to complex disease. Cell 2021;doi:10.1016/j.cell.2021.03.050.

[36] Taskinen MR. Lipoprotein lipase in diabetes. Diabetes / Metabolism Reviews 1987;3(2):551– 570. doi:10.1002/dmr.5610030208.

[37] Small KS, Todorčević M, Civelek M, El-Sayed Moustafa JS, Wang X, Simon MM, et al. Regulatory variants at KLF14 influence type 2 diabetes risk via a female-specific effect on adipocyte size and body composition. Nature Genetics 2018;50(4):572–580. doi:10.1038/s41588-018-0088-x. Number: 4 Publisher: Nature Publishing Group.

[38] Lambillotte C, Gilon P, Henquin JC. Direct glucocorticoid inhibition of insulin secretion. An in vitro study of dexamethasone effects in mouse islets. The Journal of Clinical Investigation 1997;99(3):414–423. doi:10.1172/JCI119175. Publisher: American Society for Clinical Investigation.

[39] Saad MJ, Folli F, Kahn JA, Kahn CR. Modulation of insulin receptor, insulin receptor substrate-1, and phosphatidylinositol 3-kinase in liver and muscle of dexamethasone-treated rats. The Journal of Clinical Investigation 1993;92(4):2065–2072. doi:10.1172/JCI116803. Publisher: American Society for Clinical Investigation.

[40] Vujkovic M, Ramdas S, Lorenz KM, Schneider CV, Park J, Lee KM, et al. A genome-wide association study for nonalcoholic fatty liver disease identifies novel genetic loci and trait-relevant candidate genes in the Million Veteran Program. medRxiv 2021;page 2020.12.26.20248491. doi:10.1101/2020.12.26.20248491. Publisher: Cold Spring Harbor Laboratory Press.

[41] Stender S, Kozlitina J, Nordestgaard BG, Tybjærg-Hansen A, Hobbs HH, Cohen JC. Adiposity amplifies the genetic risk of fatty liver disease conferred by multiple loci. Nature Genetics 2017;49(6):842–847. doi:10.1038/ng.3855. Number: 6 Publisher: Nature Publishing Group.

[42] Oughtred R, Rust J, Chang C, Breitkreutz BJ, Stark C, Willems A, et al. The BioGRID database: A comprehensive biomedical resource of curated protein, genetic, and chemical interactions. Protein Science 2021;30(1):187–200. doi:https://doi.org/10.1002/pro.3978. eprint: https://onlinelibrary.wiley.com/doi/pdf/10.1002/pro.3978.

[43] Thauvin-Robinet C, Auclair M, Duplomb L, Caron-Debarle M, Avila M, St-Onge J, et al. PIK3R1 mutations cause syndromic insulin resistance with lipoatrophy. American Journal of Human Genetics 2013;93(1):141–149. doi:10.1016/j.ajhg.2013.05.019.

[44] Li X, Kumar A, Zhang F, Lee C, Li Y, Tang Z, Arjunan P. VEGF-independent angiogenic pathways induced by PDGF-C. Oncotarget 2010;1(4):309–314.

[45] Folestad E, Kunath A, Wågsäter D. PDGF-C and PDGF-D signaling in vascular diseases and animal models. Molecular Aspects of Medicine 2018;62:1–11. doi:10.1016/j.mam.2018.01.005.

[46] Sansbury FH, Flanagan SE, Houghton JAL, Shuixian Shen FL, Al-Senani AMS, Habeb AM, Abdullah M, Kariminejad A, Ellard S, Hattersley AT. SLC2A2 mutations can cause neonatal diabetes, suggesting GLUT2 may have a role in human insulin secretion. Diabetologia 2012; 55(9):2381–2385. doi:10.1007/s00125-012-2595-0.

[47] Fischer TT, Ehrlich BE. Wolfram syndrome: a monogenic model for diabetes mellitus and neurodegeneration. Current Opinion in Physiology 2020;17:115–123. doi:10.1016/j.cophys.2020.07.009.

[48] Burkey BF, Hoglen NC, Inskeep P, Wyman M, Hughes TE, Vath JE. Preclinical Efficacy and Safety of the Novel Anti-diabetic, Anti-obesity MetAP2 Inhibitor, ZGN-1061. Journal of Pharmacology and Experimental Therapeutics 2018;doi:10.1124/jpet.117.246272. Publisher: American Society for Pharmacology and Experimental Therapeutics Section: Endocrine and Diabetes.

[49] Fathzadeh M, Li J, Rao A, Cook N, Chennamsetty I, Seldin M, et al. FAM13A affects body fat distribution and adipocyte function. Nature Communications 2020;11(1):1465. doi:10.1038/s41467-020-15291-z. Number: 1 Publisher: Nature Publishing Group.

[50] Hashimoto K, Shimizu E, Iyo M. Critical role of brain-derived neurotrophic factor in mood disorders. Brain Research Reviews 2004;45(2):104–114. doi:10.1016/j.brainresrev.2004.02.003.

[51] Friedel S, Horro FF, Wermter AK, Geller F, Dempfle A, Reichwald K, et al. Mutation screen of the brain derived neurotrophic factor gene (BDNF): Identification of several genetic variants and association studies in patients with obesity, eating disorders, and attention-deficit/hyperactivity disorder. American Journal of Medical Genetics Part B: Neuropsychiatric Genetics 2005;132B(1):96–99. doi:https://doi.org/10.1002/ajmg.b.30090. eprint: https://onlinelibrary.wiley.com/doi/pdf/10.1002/ajmg.b.30090.

[52] Nies VJM, Sancar G, Liu W, van Zutphen T, Struik D, Yu RT, Atkins AR, Evans RM, Jonker JW, Downes MR. Fibroblast Growth Factor Signaling in Metabolic Regulation. Frontiers in Endocrinology 2016;6. doi:10.3389/fendo.2015.00193. Publisher: Frontiers.

[53] Liu B, Gloudemans MJ, Rao AS, Ingelsson E, Montgomery SB. Abundant associations with gene expression complicate GWAS follow-up. Nature Genetics 2019;51(5):768–769. doi:10.1038/s41588-019-0404-0.

[54] Hormozdiari F, van de Bunt M, Segrè AV, Li X, Joo JWJ, Bilow M, Sul JH, Sankararaman S, Pasaniuc B, Eskin E. Colocalization of GWAS and eQTL Signals Detects Target Genes. American Journal of Human Genetics 2016;99(6):1245–1260. doi:10.1016/j.ajhg.2016.10.003.

[55] Benner C, Spencer CCA, Havulinna AS, Salomaa V, Ripatti S, Pirinen M. FINEMAP: efficient variable selection using summary data from genome-wide association studies. Bioinformatics (Oxford, England) 2016;32(10):1493–1501. doi:10.1093/bioinformatics/btw018.

[56] 1000 Genomes Project Consortium, Auton A, Brooks LD, Durbin RM, Garrison EP, Kang HM, et al. A global reference for human genetic variation. Nature 2015;526(7571):68–74. doi:10.1038/nature15393.

[57] Yu G, Wang LG, Han Y, He QY. clusterProfiler: an R package for comparing biological themes among gene clusters. Omics: A Journal of Integrative Biology 2012;16(5):284–287. doi:10.1089/omi.2011.0118.

[58] Kamburov A, Stelzl U, Lehrach H, Herwig R. The ConsensusPathDB interaction database: 2013 update. Nucleic Acids Research 2013;41(Database issue):D793–D800. doi:10.1093/nar/gks1055.

[59] [59] Csardi G, Nepusz T. The Igraph Software Package for Complex Network Research. Inter-Journal 2005;Complex Systems:1695.

[60] Bonnefond A, Froguel P. Rare and common genetic events in type 2 diabetes: what should biologists know? Cell Metabolism 2015;21(3):357–368. doi:10.1016/j.cmet.2014.12.020.

[61] Brown AE, Walker M. Genetics of Insulin Resistance and the Metabolic Syndrome. Current Cardiology Reports 2016;18. doi:10.1007/s11886-016-0755-4.

[62] Prasad RB, Groop L. Genetics of Type 2 Diabetes—Pitfalls and Possibilities. Genes 2015; 6(1):87–123. doi:10.3390/genes6010087.

[63] Melvin A, O’Rahilly S, Savage D. Genetic syndromes of severe insulin resistance. Current Opinion in Genetics & Development 2018;50:60–67. doi:10.1016/j.gde.2018.02.002.

[64] Semple RK, Savage DB, Cochran EK, Gorden P, O’Rahilly S. Genetic syndromes of severe insulin resistance. Endocrine Reviews 2011;32(4):498–514. doi:10.1210/er.2010-0020.

